# Vaccination and Non-School NPIs Shaped Ontario’s Pre-Omicron COVID-19 Hospital Burden

**DOI:** 10.64898/2026.10.14.26365553

**Authors:** Marwa Z. Tuffaha, David Dick, Jing Li, Jane M. Heffernan

## Abstract

We develop an age- and immunity-structured COVID-19 model with setting-specific contacts, calibrated to Ontario incidence and hospital admissions before Omicron. We simulate counterfactual vaccination strategies and contact restrictions in schools, workplaces, and other community settings, evaluating projected severe-case demand against an illustrative healthcare-capacity benchmark. Vaccination combined with workplace and community contact restrictions keeps simulated demand within capacity. In contrast, relaxing restrictions in either setting produces large surges and repeated capacity threats, even when school restrictions remain. School-only changes produce smaller effects on severe disease. These findings highlight the value of vaccination and setting-specific contact restrictions that prioritize workplace and community settings for pathogens with adult-concentrated severe outcomes.

## 1 Introduction

In the opening months of 2020, governments faced unprecedented choices -— act swiftly to curb transmission or risk waves of infection. With no pre-existing immunity worldwide, early projections warned that COVID-19 could cause a severe global death toll [1] and impose both shortand long-term health burdens [2]. These threats prompted the World Health Organization to issue urgent strategic guidance [3]. The delay in vaccine development forced governments to rely heavily on non-pharmaceutical interventions (NPIs)—including lockdowns, face masks, and limits on gatherings—to slow transmission by reducing human contact [4].

Although the stringency of NPIs varied across jurisdictions [4], timely mitigation measures contained infections [5], whereas delayed action overwhelmed many health-care systems [6, 7]. Nonetheless, as anticipated, these measures produced a range of enduring impacts across various sectors—such as the economy [8], education [9, 10], employment [11, 12], and mental health [13, 14].

A growing body of modelling, empirical and WHO evidence shows that SARS-CoV-2 transmission is substantially reduced by bundles of NPIs, often reducing the effective reproductive number to about 1 [5, 15]. Yet, disentangling the contribution of individual measures -— such as masking, school closure or stay-at-home orders —remains challenging. Policies are frequently implemented concurrently, and their effectiveness can depend strongly on timing, adherence, population behaviour, and baseline contact patterns [15–18].

Counterfactual modelling -— simulating alternative “what if” scenarios —is a powerful tool for estimating the potential impact of specific interventions [19–22]. In Canada, most existing studies focus on national-level analyses [23–25] or on individual NPIs in isolation [26–28], often without incorporating both age structure and immunity dynamics (from infection and vaccination). When age structure is included, the analysis is often performed using stochastic simulation models [24, 29, 30] (but see [31]), which, while detailed, are computationally intensive and slow to run.

Here, we apply a modified Age-Structured Immunity and Infection Model (ASIIM) [32, 33] to evaluate counterfactual NPI scenarios in Ontario, Canada. Ontario provides a useful case study because it is Canada’s most populous province and implemented province-specific NPIs. Unlike previous Ontario studies [21, 27, 28, 34], our computationally efficient ordinary differential equation (ODE) framework incorporates age structure, varying immunity levels, and vaccination. It enables us to assess not only the combined effects of vaccination and NPIs, but also whether changes in school, workplace, and other community contacts differentially affect severe disease and hospital demand. We restrict the analysis to the pre-Omicron period, allowing us to examine how the timing and stringency of vaccination and NPIs shape projected outcomes.

## 2 Methods

### 2.1 Model Structure

We extend the ASIIM model from previous work [32, 33] to better track COVID-19 vaccine history and status within a population. The full model equations and compartment transitions, including vaccination, infection progression, recovery, and immunity waning, are detailed in Supplementary Section S1 and illustrated in Supplementary Figure S1.

Briefly, the model tracks disease and immunity status, as well as infection severity by age. Susceptible, exposed, and infected populations are split into vaccinated and unvaccinated subgroups. Immunity strength is divided into four discrete levels (1–4, from low to high immunity), and infection symptoms and infectiousness characteristics are tracked over three severities of infection (2 = mild/asymptomatic, 3 = moderate/some symptoms, 4 = severe/symptomatic and reported to healthcare). The population is split into 16 age groups in 5-year blocks, enabling age-specific contact transmission, disease progression, and vaccination schedules. This stratification allows the model to represent eligibility for first, second, and booster doses, while also accounting for waning immunity, and agespecific contact behaviour. Although demographic dynamics are not active in the present analysis, they are included in the model equations and parameter definitions provided in the Supplement and may be used in future applications.

Age-specific transmission is driven by contact matrices for household, school, workplace, and other community settings. These matrices are modified over time to reflect non-pharmaceutical interventions (NPIs), using the school, workplace, and community restriction levels reported in the Canadian Provincial COVID-19 Tracker (CPRCT) dataset [35]. In this way, changes in public health restrictions are translated into time-varying contact patterns between age groups. The age-specific baseline contact patterns and the setting-specific multipliers used to represent different NPI levels are illustrated in Supplementary Figures S2 and S3, respectively.

Within each vaccination-status and immunity level, individuals move through susceptible, exposed (multi-stage latent period – infected but not infectious), and infected (infectious) compartments. Severity outcomes are determined through age-specific probabilities based on potential comorbidities [36]. Vaccination has a failure rate, but when effective, it decreases the risk of infection by moving individuals to higher levels of immunity and decreases the risk of severe disease by modifying the age-specific probabilities to reflect vaccine effectiveness in mitigating symptoms.

### 2.2 Data

We restrict our study to the pre-Omicron phase of the COVID-19 pandemic for three reasons. First, NPI and vaccination policies were implemented mainly in the pre-Omicron phase of the pandemic. Second, because the Omicron variant had significantly different properties from earlier variants [37], the model assumptions may not align with reality. Lastly, government data collection and testing efforts were greatly reduced starting from the Omicron phase [38].

We use daily incidence (public surveillance) data from Berry et al. [39]. Vaccination coverage data for Ontario are from the Public Health Agency of Canada COVID-19 vaccination coverage dataset [40]. Daily hospitalization data is taken from Betti et al. [41]. Changes in public health mitigation and their effects of contact structure within the population are taken from Akanteva et al. [35].

#### 2.2.1 Calibration to Data

The mathematical model is calibrated to daily incidence data, to daily hospital admission data, or to both simultaneously. We calibrate the force of infection, *λ*(*t*), over time using a time-varying multiplier, *κ*(*t*). We note that *λ*(*t*) depends on age-structured population mixing, infection prevalence, and the infectivity of each severity class (see Supplementary Information for details). The infectivity parameters within the force of infection parameter are selected such that the initial force of infection, *λ*(0), corresponds to a basic reproductive number within the observed range for COVID-19 in Ontario (*R*_0_ = 2.6) [42]. To capture variability in public health policy regarding masking and social distancing, we implement public health mitigation through modifications on contract structure within the population [35]. To capture variability on human behaviour (e.g. compliance with NPIs) and changes in infectivity associated with emerging variants, we scale the force of infection by time-varying parameter *κ*(*t*), which is calibrated to the observed hospitalization and infection incidence data.

The parameter *κ*(*t*) acts as a time-varying multiplier on transmission that captures changes not fully represented by the age-structured contact matrices alone. Thus, values above unity indicate higher effective transmission than predicted by the baseline contact structure, which may reflect more relaxed behaviour, reduced adherence to masking or distancing, increased transmissibility of circulating variants, seasonal/weather-related effects, or other unmodelled transmission factors. Conversely, values below unity indicate lower effective transmission than predicted by the contact matrices, consistent with stronger adherence to protective behaviours or other factors reducing transmission.

The model is calibrated in MATLAB using a genetic algorithm to estimate weekly values of *κ*(*t*) that minimize the percent error between observed data and model predictions. The calibration objective is evaluated against LOESS-smoothed observed time series. LOESS, or locally estimated scatterplot smoothing, is a nonparametric local-regression method in which the smoothed value at each time point is obtained from a weighted low-degree polynomial regression on nearby observations, with observations closer to the target time point receiving greater weight [43]. This reduces the influence of short-term reporting noise while preserving the main epidemic trends used for calibration.

To compare the model with surveillance data, we derive daily incidence from the flow of individuals progressing from the final exposed stage into the corresponding infectious compartments. We assume that all incident severe infections are detected, that 60% of incident moderate infections are detected, and that mild infections are not systematically detected. Daily hospital admissions are estimated as 15% of daily severe incidence. These assumptions allow the model outputs to be translated into quantities comparable to the reported daily case counts and daily hospital admissions. We assess the sensitivity of the calibration to these assumptions by varying the detection and hospitalization proportions (see Section 2.2.4).

When the model is calibrated to both daily cases and daily hospital admissions, we measure the error between the model output and the data as a percent error rather than as an absolute difference. This prevents the larger case counts from dominating the calibration and gives the hospitalization data comparable influence on the estimated parameters. In this joint calibration, the objective function is therefore defined as the sum of the percent error for daily cases and the percent error for daily hospital admissions.

Rather than estimating all weekly values of *κ*(*t*) simultaneously, we use a 16-week sliding calibration window to balance local accuracy and computational cost. We initialize *κ*(*t*) = 1 for all weeks, representing no variability in human behaviour or viral properties, and allow the algorithm to search for weekly values of *κ*(*t*). Typically, *κ*(*t*) rises above 1 when the uncalibrated model underpredicts infections and falls below 1 when it overpredicts them.

Because the genetic algorithm is stochastic, the calibrated values of *κ*(*t*) are not unique and may vary between runs. However, the overall model dynamics remain consistent. This is because small deviations in *κ*(*t*) at a particular week can be compensated by adjustments in subsequent weeks, so that what matters is the aggregate effect of *κ*(*t*) over a given time interval rather than the exact week-to-week values. Given this small variation between calibrations, we perform 10 independent calibration runs and report model outputs averaged across these runs. For visualization of the calibrated transmission multiplier itself, Figure 3 shows the mean *κ*(*t*) across the 10 runs together with the variation among runs. To further promote stability and avoid unrealistic fluctuations, we apply a curvature penalty in the objective function (set at 10% of the objective value), which discourages abrupt changes in consecutive *κ* values while still allowing sufficient flexibility to capture genuine shifts in transmission dynamics.

#### 2.2.2 Comparison to Seroprevalence Data

We compare model immunity outputs with Canadian Blood Services (CBS) seroprevalence estimates of anti-nucleocapsid (anti-N) and anti-spike (anti-S) antibodies [44]. Anti-N reflects infection-associated antibody exposure, whereas anti-S reflects exposure through infection or vaccination. Because the model’s immunity classes represent broader protection—including cell-mediated and innate components—rather than antibody concentrations alone, this comparison is intended as an external qualitative check rather than a direct validation of antibody dynamics. As blood donors are aged 17 years and older, we report model immunity for ages 15 years and above. Anti-S is compared with compartments reflecting prior spike exposure through infection or vaccination; anti-N is compared with lower and upper model-based bounds reflecting prior infection because vaccinated recovered compartments may contain mixed vaccination and infection histories. The specific model compartments and bounds used for these comparisons are described in Supplementary Section S1.4.

### 2.3 Comparison to Vaccination Data

We compare model-predicted vaccination uptake with reported Ontario vaccination coverage data from the Public Health Agency of Canada. For first and second doses, the reported proportion of the population vaccinated is compared with the corresponding cumulative model vaccination compartments. Model outputs are averaged across the 10 independent calibrations and expressed as a proportion of the model population. This comparison provides an additional check that the vaccination dynamics implemented in the model reproduce the observed timing and overall magnitude of Ontario’s vaccine rollout.

#### 2.2.4 Sensitivity Analysis

We conduct sensitivity analyses to assess the robustness of model calibration and counterfactual results to uncertainty in infection detection and hospitalization proportions, school contact perturbations, the initial infected population, and immunity waning. These assumptions can affect inferred transmission dynamics and counterfactual predictions because surveillance, contact patterns, epidemic seeding, and immunity duration are uncertain. First, we evaluate alternative calibration parameter sets by varying the assumed detection proportions for mild, moderate, and severe infections, as well as the proportion of severe infections resulting in hospital admission. For each parameter set, we recalibrate the model to observed incidence and hospital-admission data and compare the calibration errors for each outcome and their combined error to identify the parameter set providing the most balanced calibration across both datasets (Supplementary Section S2.3.1). Second, we test alternative school contact perturbation matrices representing different reductions in school contacts under partial mitigation measures. For each matrix, we repeat the school counterfactual simulations and compare hospital prevalence trajectories to assess the sensitivity of the estimated effects of school restrictions to the assumed effectiveness of mitigation measures (Supplementary Section S2.3.2). Third, we vary the assumed initial infected population, *I*_0_, and recalibrate the model to examine its effect on the inferred transmission multiplier *κ*(*t*) and the resulting counterfactual trajectories (Supplementary Section S2.3.3). Finally, we vary the immunity waning rate, *ω*, over a range corresponding to average immunity durations of 3 months to 3years, compared with the baseline assumption of 1 year. For each waning rate, we recalibrate the model and examine changes in *κ*(*t*) and counterfactual hospital prevalence to assess the sensitivity of the predicted outcomes to immunity duration (Supplementary Section S2.3.4).

### 2.3 Counterfactual Scenario Design

Counterfactual scenarios vary vaccination and setting-specific NPIs in school, workplace, and other community settings. Figure 1 outlines all scenarios considered. We refer to the calibrated baseline scenario, which preserves NPIs and vaccination rollout as observed, as the Observed scenario. This serves as the baseline for all counterfactual comparisons. We group the counterfactual analyses into two sets: *Vaccine–NPI Crossovers*, which disentangle the respective roles of vaccination and NPIs in reducing infections, and *Targeted NPI Adjustments*, which assess the contribution of measures in individual settings.

**Figure 1:**
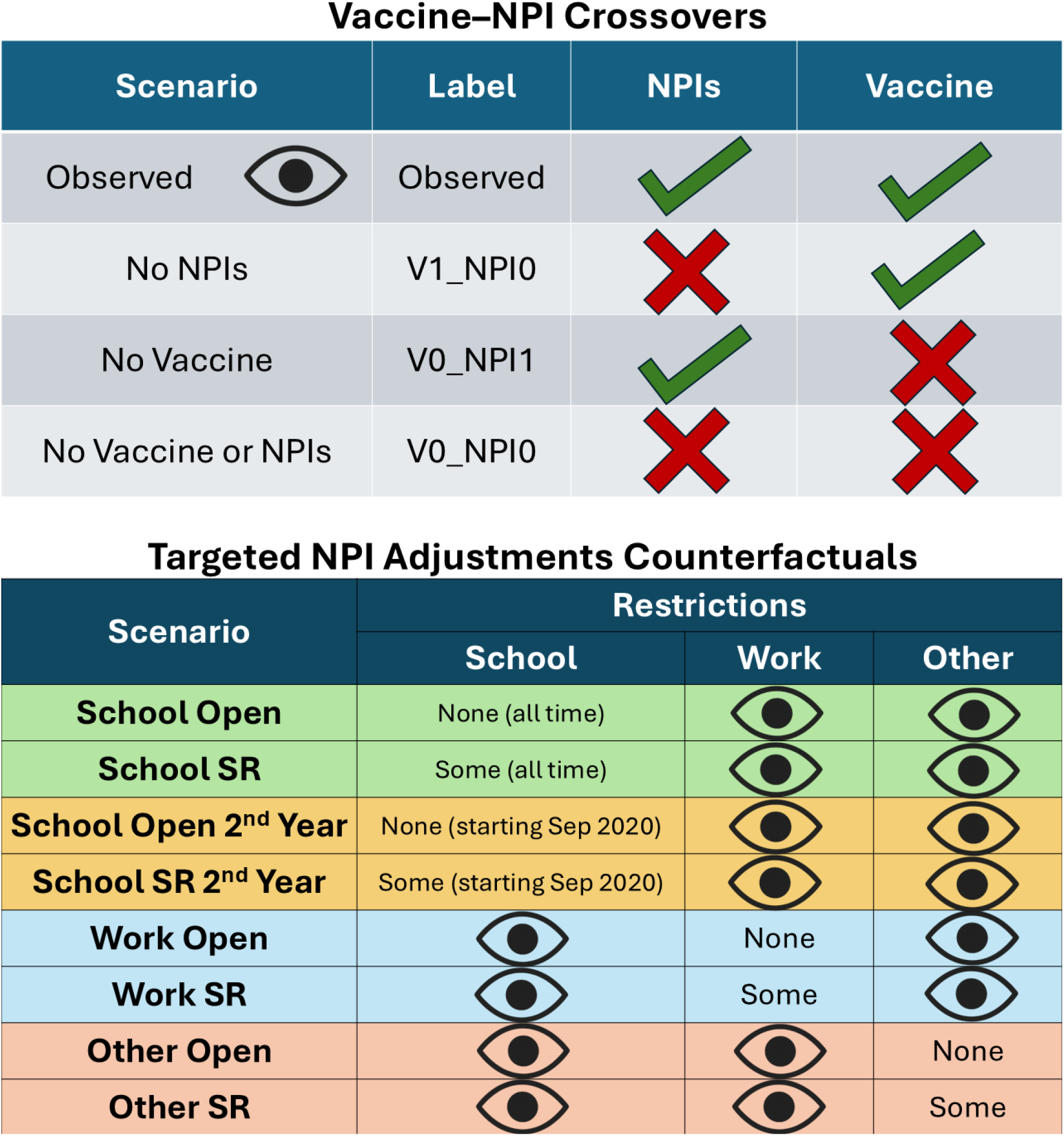
Overview of counterfactual scenarios. The upper table defines the vaccine–NPI crossover scenarios; the lower table defines targeted NPI adjustments in school, workplace, and other settings, with unchanged settings kept at their observed values.

#### 2.3.1 Vaccine–NPI Crossovers

We define three counterfactual vaccine–NPI crossover scenarios to isolate the effects of vaccination and NPIs. First, we consider no vaccination or NPIs (`V0_NPI0`) to quantify outcomes in the absence of both public health interventions. We then consider NPIs only (no vaccination; V0 NPI1) and vaccination only (no NPIs; V1 NPI0). Comparing V0 NPI1 with `V0_NPI0` estimates the effect of NPIs in a vaccine-free context, whereas comparing V1 NPI0 with `V0_NPI0` estimates the effect of vaccination in the absence of NPIs.

#### 2.3.2 Targeted NPI Adjustments

Here, we compare setting-specific NPI changes with the Observed scenario to quantify the effects of school, workplace, and other-setting restrictions on COVID-19 healthcare demand and infection outcomes. We construct counterfactuals that modify NPIs in a single setting while holding vaccination and all other controls at their observed values. As shown in Figure 1, we consider no restrictions throughout the study period (*Open*), some restrictions throughout the study period (*SR*), and, for schools, scenarios in which either no restrictions or some restrictions are applied beginning in September 2020 (*School Open 2nd Year* and *School SR 2nd Year*, respectively). Before September 2020, these latter school scenarios retain the observed school restrictions. In each case, only the named setting is altered.

We use “some restrictions” (*SR*) to refer to Level 1 restrictions in the CPRCT dataset [35]: distancing, masking, and enhanced cleaning in schools; recommended work-from-home or businesses operating with some restrictions in workplaces; and minor gathering limits and related measures in other community settings. The scenarios allow us to compare each counterfactual with the Observed trajectory and to assess how the restriction level and timing affect the resulting epidemic trajectory.

#### 2.3.3 Comparing Counterfactual Results

We compare counterfactual scenarios in terms of acute-care demand by estimating simulated hospital prevalence from model-predicted hospital admissions. In the model, hospital admissions are estimated as 15% of daily severe incidence, which is obtained from the flow of individuals progressing from the final exposed stage into the severe infectious compartments. The baseline proportion of 15% is selected from the calibration-parameter comparison described above. To convert daily admissions into daily hospital occupancy, we assume that hospital length of stay follows a lognormal distribution with median 8.7 days and interquartile range 3.6–18.9 days [45]. The simulated admissions time series is then convolved with the corresponding length-of-stay survival probabilities to estimate the number of patients still hospitalized on each day.

For visual context, we overlay Ontario’s nominal acute-care capacity: 29,743 ward beds and 2,300 ICU beds [41]. These reference lines indicate potential pressure rather than literal overload thresholds because not all beds are available for COVID-19 care and staffed capacity may be substantially lower during surges; crossings should therefore be interpreted as indicative rather than literal exceedances.

## 3 Results

### 3.1 Model Calibration and the Observed Scenario

When calibrated to incidence or hospital admissions alone, the model more closely reproduces the corresponding data stream but shows larger discrepancies in the other outcome (Supplementary Figures S4 and S5). Simultaneous calibration captures the main features of both observed trajectories, including the Alpha and Delta waves and the decline following vaccine rollout (Figure 2, top panels), while also reproducing their distinct age distributions (Figure 2, bottom panels). Joint calibration has the lowest combined calibration error, whereas single-target calibration minimizes the error for its respective outcome (Supplementary Figure S6).

**Figure 2:**
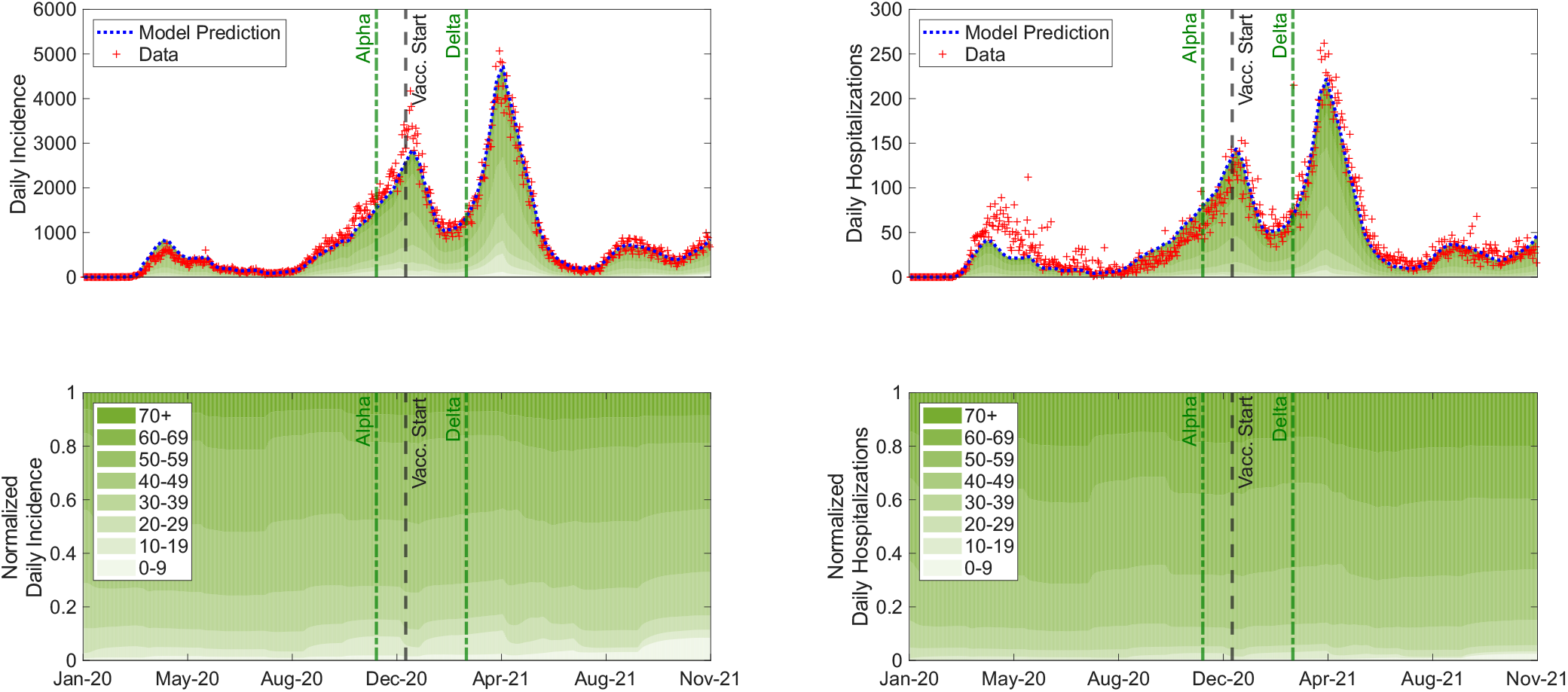
Model calibration to observed Ontario daily incidence and hospital admissions. Top panels compare model predictions with surveillance data; bottom panels show the normalized age distributions of incidence and hospital admissions.

The weekly *κ*(*t*) profiles from joint calibration are shown in Figure 3. Values rise above 2 early in the epidemic, suggesting that the contact matrices alone are too restrictive to reproduce the first wave. Values close to 1 are then sufficient through the Delta wave, whereas values above 3 are required after the Delta peak to reproduce the sustained plateau in cases and hospitalizations. This pattern suggests additional transmission pressures beyond the baseline contact structure, including behavioural relaxation, variant effects, seasonal factors, or other unmodelled changes.

**Figure 3:**
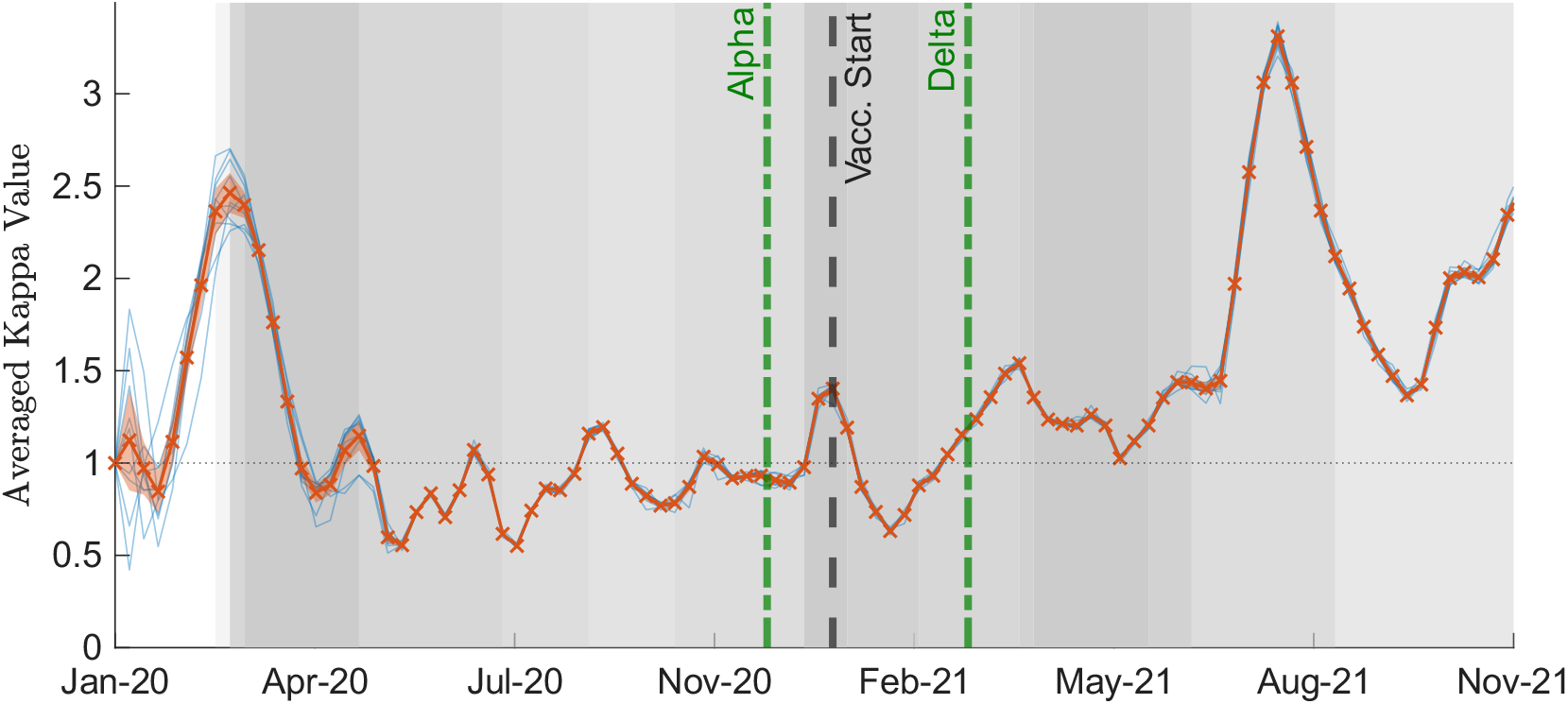
Calibrated transmission multiplier *κ*(*t*) from the joint calibration to incidence and hospitalization data. Faint blue curves show 10 independent calibration runs; the red curve shows the mean with 95% confidence intervals. Grey shading indicates the overall strictness of NPIs over time, with darker shading corresponding to stricter measures.

Importantly, the calibration captures the distinct age distributions of incidence and hospitalizations (Figure 2, lower panels): infections are spread broadly across younger and middle-aged groups, particularly ages 20–49, whereas hospital admissions are heavily concentrated among older adults aged 50 and above, consistent with provincial surveillance data. In addition, the model reproduces the observed vaccination rollout and broad population-level immunity trends (Figure 4). The cumulative first- and second-dose model compartments closely follow the corresponding reported vaccination coverage (Figure 4, top). For the seroprevalence comparison (Figure 4, bottom), the simulated anti-S profile closely follows the observed trend across the study period, whereas anti-N prevalence is slightly underestimated early on but later rises above the lower bounds of the data. Because the model tracks overall immunity rather than directly measuring seroprevalence, we do not expect perfect agreement with antibody-based estimates. These differences suggest some mismatch in how the model translates infections into detectable antibody responses; we return to their implications for interpreting infectionversus vaccine-derived immunity in the Discussion. Together, these results demonstrate that the calibration procedure not only reconstructs epidemic trajectories, but also captures the broad age-specific disease burden and population-level immunity patterns.

**Figure 4:**
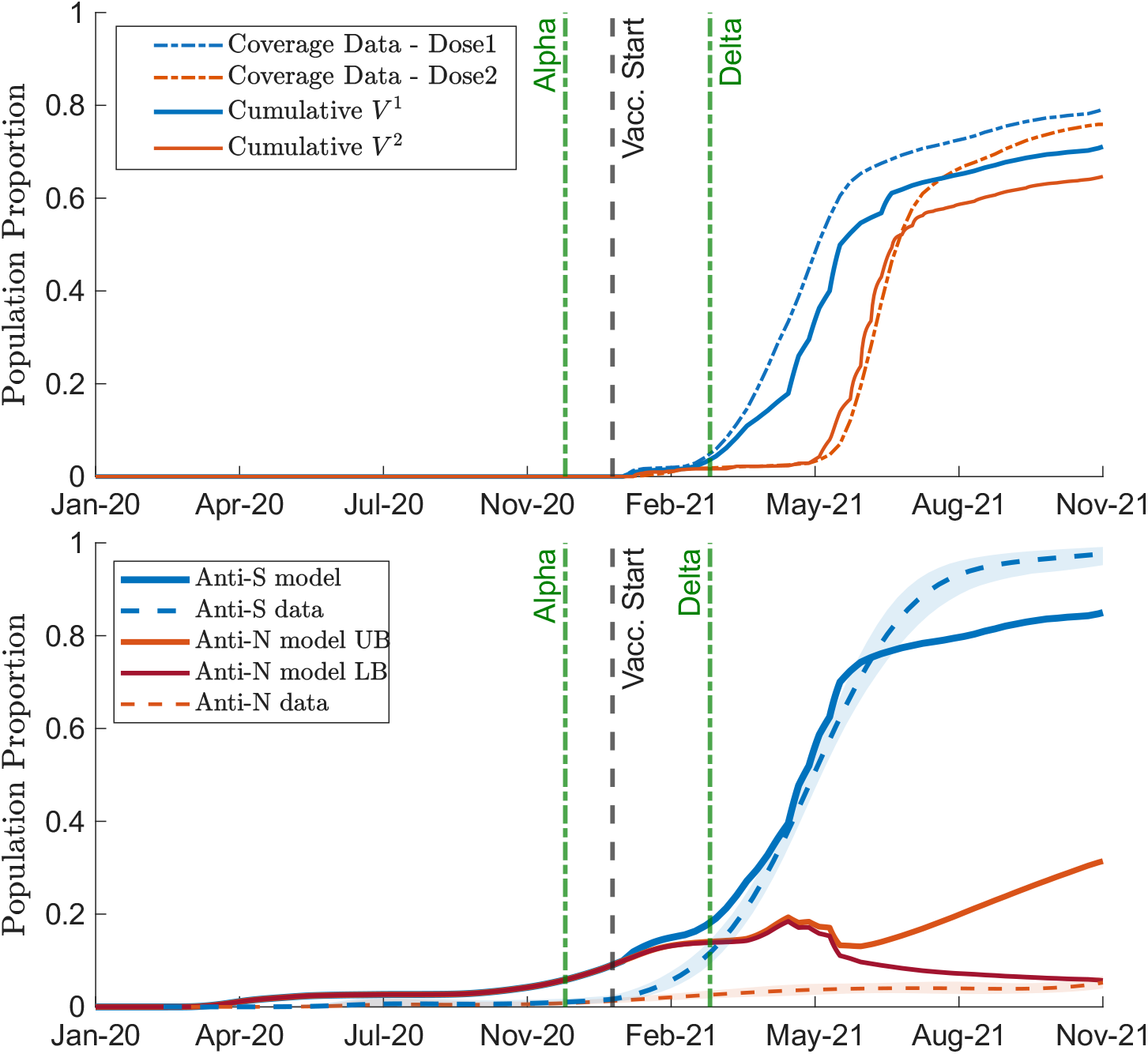
Comparison of model vaccination and immunity outputs with observed data. Top: Comparison of reported Ontario first- and second-dose vaccination coverage with the corresponding cumulative model vaccination compartments. Dashed curves show reported vaccination coverage and solid curves show model outputs averaged across the 10 calibrations. Bottom: Comparison of model-derived immunity measures with seroprevalence data for ages 15 years and older. Anti-S reflects prior spike exposure from infection or vaccination, while anti-N is shown using lower and upper model-based bounds because vaccination and infection histories are partly mixed within the model compartments.

Under the Observed scenario, estimated hospital prevalence follows the successive epidemic waves, including the Alpha and Delta periods, while remaining below 25% of Ontario’s acute-care capacity throughout the study period (Figure 5, blue curves). The corresponding trajectories of mild (*I*_2_), moderate (*I*_3_), and severe (*I*_4_) infections exhibit similar temporal patterns, with differences in magnitude across severity classes (Supplementary Figures S7–S9, blue curves). Hospital prevalence follows the general pattern of severe infections, with a smoother trajectory reflecting the duration of hospital stays. The immunity distribution for each age class is shown in Figure 7 (top panel). Under the Observed scenario, most of the population remains at the lowest immunity level throughout 2020. Following vaccine rollout, immunity accumulates primarily through vaccination, shifting the population toward higher immunity levels. By the end of 2021, nearly half of the population reaches immunity level 4, while approximately one-quarter remains at level 1, predominantly among the youngest age groups.

**Figure 5:**
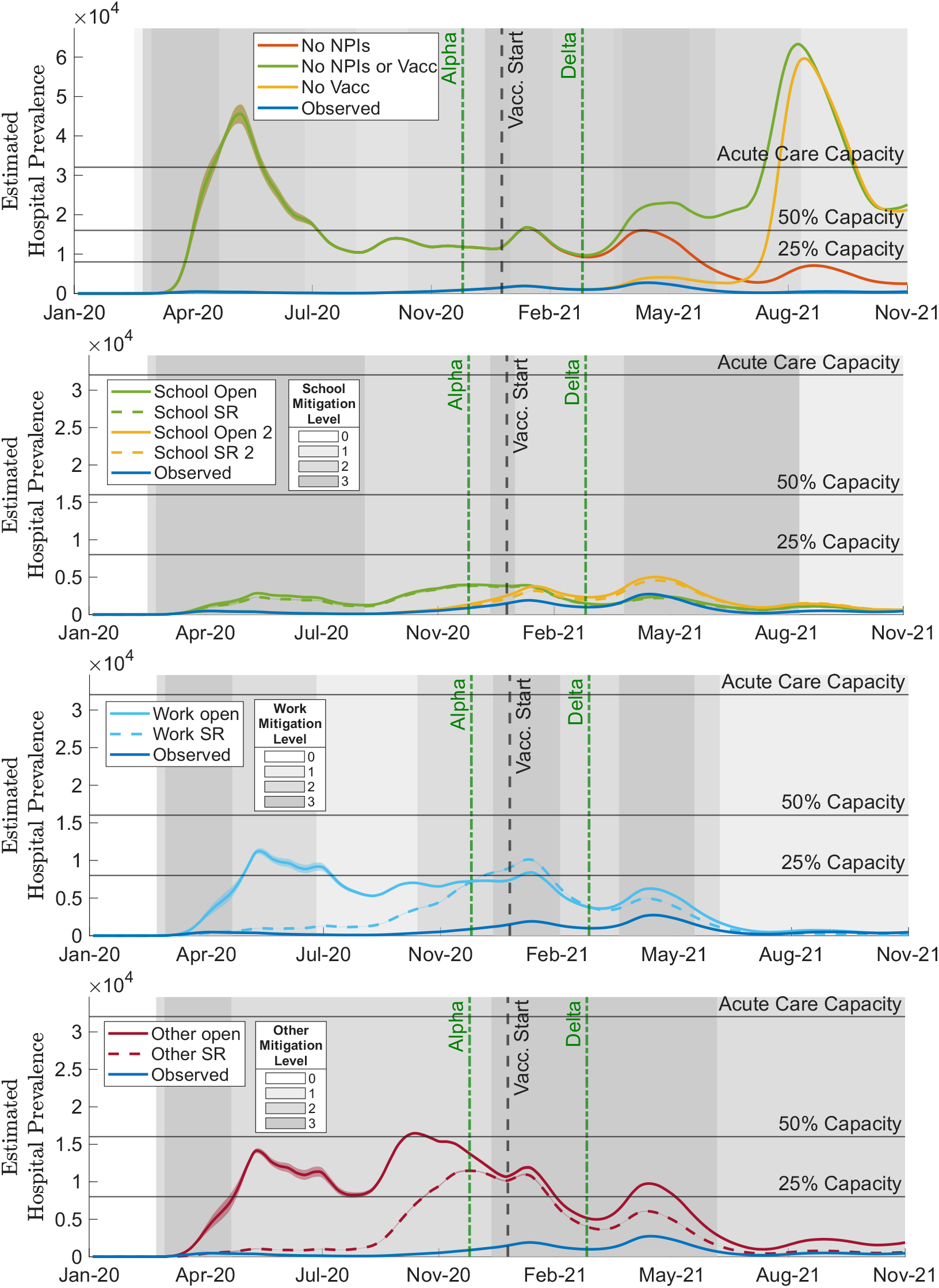
Estimated hospital prevalence under counterfactual scenarios. Panels show vaccine–NPI crossovers, followed by targeted changes to school, workplace, and other-setting NPIs. Horizontal lines indicate 25%, 50%, and 100% of Ontario acute-care capacity. Grey shading is based on the NPI levels in the observed scenario and is included for timing context: the first panel shows the overall aggregate NPI level, while the remaining panels show the observed setting-specific NPI level for the corresponding school, workplace, or other setting. Darker shading indicates stricter measures.

Sensitivity analyses indicate that the model results and conclusions are robust to the modelling assumptions examined. Alternative detection and hospitalization proportions show the same trade-off, supporting the joint calibration choice (Supplementary Table S5; Supplementary Figure S10). Stronger assumed effectiveness of school-contact reductions has little impact on hospital prevalence under the Observed scenario (Supplementary Figure S11, blue curves). Varying the initial infected population primarily affects the inferred *κ*(*t*) during the early epidemic, with differences diminishing thereafter (Supplementary Figure S12). Similarly, faster immunity waning increases projected hospital prevalence without substantially altering the relative patterns across counterfactuals (Supplementary Figure S13).

The calibrated model compartment trajectories illustrate the underlying immunity dynamics, including the progression from first- to second-dose vaccination following rollout and the subsequent accumulation of vaccinated-susceptible individuals as immunity wanes (Supplementary Figure S14).

### 3.2 Counterfactual Comparisons

#### 3.2.1 Vaccine-NPI Crossovers

Considering vaccine–NPI crossovers (Figure 5, top panel), removing both vaccination and NPIs (`V0_NPI0`, green) produces an early, explosive peak in hospital prevalence well above acute-care capacity. Hospital prevalence then declines to approximately 50% of capacity before rising above capacity again in the post-Delta period. With NPIs but no vaccination (V0 NPI1, yellow), the first large surge is delayed until the post-Delta period, but it remains similar in magnitude to the corresponding `V0_NPI0` surge and substantially exceeds acute-care capacity. Vaccination without NPIs (V1 NPI0, red) also produces hospital prevalence above acute-care capacity, although it falls below this threshold during short periods in 2021. Only the Observed combination of vaccination and NPIs maintains hospital prevalence below 25% of acute-care capacity throughout the study period.

The corresponding trajectories of mild, moderate, and severe infections show consistent patterns: removing NPIs produces large early infection waves, whereas removing vaccination increases the later epidemic burden (Supplementary Figures S7–S9).

#### 3.2.2 Targeted NPI Adjustments

Targeted NPIs reveal setting-specific effects (Figure 5). In the school scenarios (second panel), keeping schools fully open throughout both academic years leads to a substantial increase in hospital prevalence, with some peaks reaching more than twice those of the observed trajectory. Applying some restrictions (SR) in schools makes little difference. By contrast, opening schools only in the second academic year (yellow curves) shifts the burden to the post-Alpha and post-Delta periods. All school scenarios result in hospital prevalence below 25% of acute-care capacity.

For workplaces (Figure 5, third panel), full reopening (light blue) drives hospital prevalence up well above and then around the 25% of acute-care capacity line until vaccination is applied. With some workplace restrictions (Work SR, light blue dashed line) the initial peak is delayed until the post-Alpha period, but hospital occupancy is curbed by vaccination and there is only a relatively small surge of infection in the post-Delta period. However, levels still remain clearly above the Observed scenario.

In the “other” setting (gatherings/venues), relaxing measures produces the largest single-setting increase, hitting the 50% occupancy line, with SR blunting but not eliminating the surge (Figure 5, bottom panel). We observe levels well-above the 25% of acute-care capacity line, with decreases below this threshold only after vaccination is initiated though it is briefly passed again during the Delta wave.

Overall, NPIs shape both the timing and magnitude of hospital burden. V0 NPI1 delays the first large wave but does not reduce it enough to avoid capacity breaches. Among targeted policies, relaxing restrictions in other settings produces the largest increase in hospital prevalence, followed by workplace reopening and then school reopening. Only the Observed strategy and the school-only counterfactuals maintain hospital prevalence below 25% of acute-care capacity throughout the study period.

The infection trajectories confirm that these setting-specific differences extend across all three severity classes, with community and workplace restrictions producing substantially greater reductions in mild, moderate, and severe infections than school-focused measures (Supplementary Figures S7–S9).

Because the timing and number of peaks differ across scenarios, we also summarize hospital prevalence within fixed wave windows using the wave maximum and the peak 6-week mean (Supplementary Figure S15; Supplementary Section S2.5). This wave-specific summary shows that the largest burdens occur in different periods for different counterfactuals: removing NPIs produces an immediate first-wave capacity threat, removing vaccination mainly amplifies the post-Delta period, and relaxing workplace or other-setting measures creates large pre-vaccine and Alpha-period burdens. These results reinforce that vaccination and layered NPIs jointly shape both the timing and magnitude of hospital demand.

Cumulative outcomes show the same pattern over the full study period (Figure 6). Removing both vaccination and NPIs produces the largest increases in total infections and hospital admissions, and removing either intervention alone also substantially increases burden. Among targeted adjustments, relaxing other-setting measures has the largest cumulative effect, followed by workplace measures, whereas school changes have considerably smaller effects. Within each setting, retaining some restrictions reduces cumulative burden, and opening schools only in the second academic year reduces excess burden relative to opening throughout both years. This pattern is consistent across mild, moderate, and severe infections (Supplementary Figure S16).

**Figure 6:**
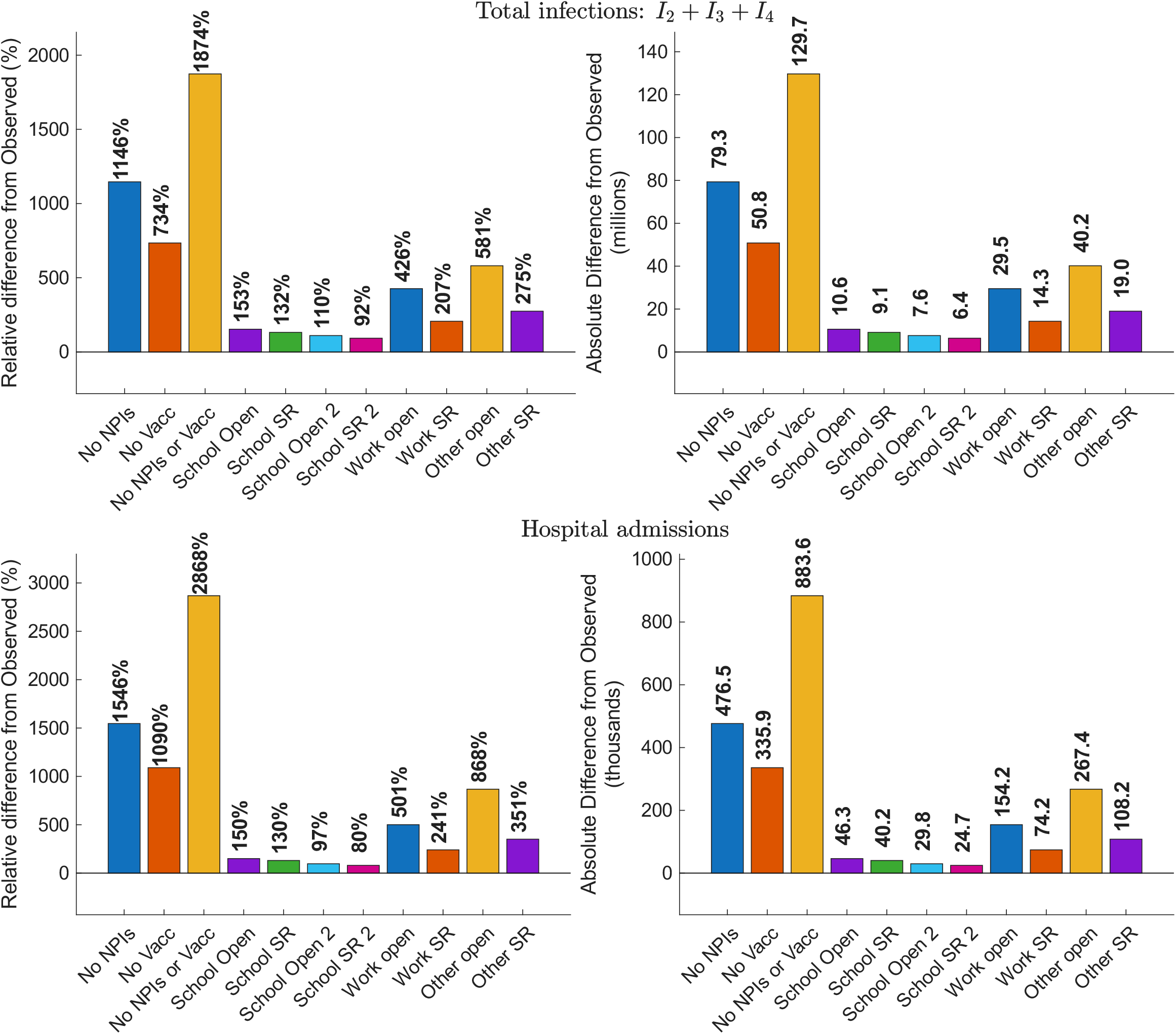
Differences in cumulative total infections and hospital admissions compared with the Observed scenario. The top row shows cumulative total infections, calculated from the daily flow of individuals progressing from the final exposed stage into infectious compartments, and the bottom row shows cumulative hospital admissions. In each row, the left panel shows the relative difference from the Observed scenario, and the right panel shows the absolute difference.

### 3.3 Immunity Distributions

Figure 7 illustrates the evolution of immunity distributions across age groups and immunity levels (1–4) under the Observed scenario (top), alongside two contrasting counterfactuals: `V0_NPI0` (middle), which produces the greatest disease burden, and School Open (bottom), which has the smallest impact among the full-opening targeted NPI scenarios.

**Figure 7:**
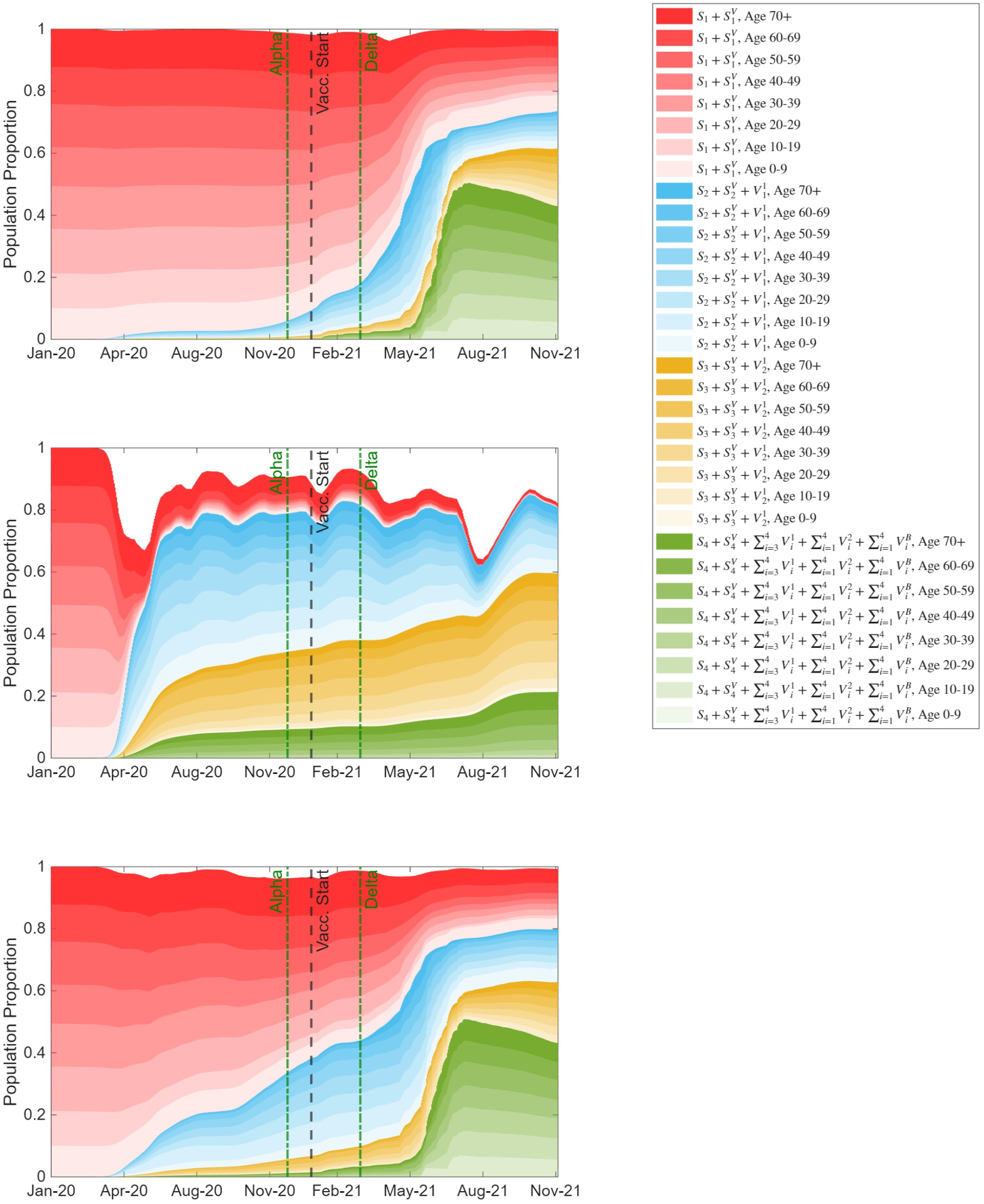
Immunity distribution by age group and immunity class in the observed, `V0_NPI0`, and School Open scenarios. Colours indicate immunity level: red represents immunity level 1, blue level 2, yellow level 3, and green level 4; shades within each colour indicate age groups. White space at the top of each panel represents exposed and infectious individuals.

In `V0_NPI0` (middle; no vaccination, no NPIs), immunity is gained almost exclusively—and very quickly—through infection, which is represented by the white area at the top of the figure. The infected fraction rises to ∼ 40% in April 2020, after which accumulated infection-derived immunity is associated with prevalence remaining in the ∼ 6–18% range until a post-Delta surge pushes infections back to ∼ 40%. By then, the naïve fraction is very small and almost entirely in the elderly (dark red). Younger groups, with more frequent exposures, move rapidly into higher immunity strata, while older adults lag and remain vulnerable early on. The composition by severity is also distinct: younger people are more visible in immunity class 2 (reflecting mild infections), whereas adults and the elderly concentrate in classes 3–4 (reflecting more severe infections).

The School Open scenario (bottom) shows an intermediate pace of immunity buildup in 2020 between the two extremes: by the vaccine start date, ∼ 40% of the population has some immunity, with the infected fraction (white area at the top) staying < 5% throughout. Subsequent vaccination then lifts overall immunity in a pattern without clear age stratification, yielding curves that resemble the observed scenario post-rollout. In short, school opening modestly accelerates pre-vaccine immunity accumulation, but once vaccines arrive, the overall and age-specific distributions converge toward the Observed scenario.

The additional counterfactuals support these observations. Without NPIs, substantial infection-derived immunity accumulates before vaccination, whereas retaining NPIs delays this accumulation in the absence of vaccination (Supplementary Figure S17). Similarly, opening workplaces and other settings accelerates pre-vaccine immunity accumulation, particularly in the Other Open scenario, while subsequent vaccination shifts the population toward higher immunity levels (Supplementary Figure S18). Overall, these results highlight that similar levels of population immunity can arise through markedly different pathways: vaccination enables immunity accumulation while limiting infection burden, whereas relaxing NPIs accelerates infection-derived immunity at the cost of greater disease burden.

## 4. Discussion

Our counterfactuals show clear setting-specific effects on epidemic peaks and cumulative disease burden. Relaxing measures in community (“other”) venues produces the largest surges in severe infections and cumulative increases in total infections and hospital admissions, followed by workplace reopening (Figure 6). Partial restrictions reduce, but do not prevent, substantial additional burden. School policies have smaller effects, although timing matters: opening throughout both academic years roughly doubles severe cases relative to the Observed scenario, whereas opening only in the second year halves and shifts the excess burden to the post-Delta period. Thus, workplace and community controls have greater influence on severe outcomes than school measures, and layered vaccination and NPIs are crucial.

Evidence on NPI effectiveness is mixed by phase and outcome. A first-wave systematic review concludes that school closures are the most effective NPI, followed by workplace and venue closures [18]. Cross-country modelling likewise finds large effects for limiting gatherings and closing high-exposure settings, including schools [16, 17]. However, some Ontario-focused studies indicate the population-level impact of school policies on older adults—and thus on severe outcomes—is limited: school closures alone change community incidence only modestly [21, 46], and reopening schools produces comparatively few infections among people > 60 years [47]. A rapid review by El Jaouhari et al. [48] finds that many studies around the globe find that school closures have limited impact on COVID-19 transmission. These patterns align with our finding that restrictions in workplaces and community venues carry greater leverage over severe disease. Given the substantial educational costs entailed by school closures [27], school-targeted NPIs may warrant cautious re-evaluation for COVID-19-like diseases, particularly after the first pandemic academic year. By that time, evidence increasingly indicated that severe COVID-19 outcomes were concentrated in older adults [49, 50], while prolonged disruption to in-person learning carried substantial educational, social, and mental-health costs [51, 52]. In our model, implementing “some restrictions” in schools (e.g., masking, distancing, and hygiene measures) yields only a modest reduction in severe outcomes compared with full opening (Figure 5, second panel). To assess whether this relatively small effect reflects the assumed modest reduction in school contact rates (Supplementary Figure S3), we repeat the school counterfactual analyses using progressively greater reductions in school contacts (Supplementary Section S2.3.2; Supplementary Figure S11). Stronger assumed mitigation increases the separation between the School Open and School SR counterfactuals, but school-related hospital prevalence remains comparatively low across all tested assumptions. These results indicate that the limited effect of school interventions on severe outcomes cannot be explained solely by the assumed magnitude of school contact reductions.

Taken together, these findings suggest that, for pathogens with adult-concentrated severe outcomes, school-focused restrictions may yield smaller marginal reductions in severe disease than measures targeting adult-dense settings while imposing substantial educational and social costs. Ontario nevertheless experienced among the longest periods of school disruption in Canada [53], reflecting concerns about infection risk, staffing, isolation requirements, and parental work disruption. This interpretation is consistent with evidence that mitigated school reopening was not generally associated with large increases in community transmission, that resurgent transmission in 2020–2021 was sustained primarily by adults aged 20–49, and that the effect of school measures depends on the wider community contact landscape [54–56]. School policy must still account for heterogeneous paediatric risk: medically vulnerable and immunocompromised students may require layered protections and reasonable accommodations, including ventilation and filtration, high-filtration masks during periods of elevated transmission, test-to-stay approaches, or short-term remote access [57, 58].

Interestingly, the calibrated *κ*(*t*) values provide insight into temporal variation that is not explicitly represented elsewhere in the model. The different calibration runs under the same parameterization yield broadly similar *κ*(*t*) trajectories (Figure 3), with the greatest variability occurring at the beginning of the epidemic and during several later transition periods. Because *κ*(*t*) acts as a time-varying transmission multiplier, these fluctuations should be interpreted as a residual signal that can absorb changes in transmission, behaviour, reporting, viral characteristics, and other processes not explicitly captured by the model, rather than as a direct measure of any single mechanism. The sensitivity analyses help distinguish fluctuations in *κ*(*t*) that arise from uncertain model assumptions from those that persist across alternative assumptions. Varying the initial infected population changes *κ*(*t*) primarily during the first few weeks of the epidemic, with the resulting trajectories rapidly converging thereafter (Supplementary Section S2.3.3; Supplementary Figure S12). Thus, the later temporal structure in *κ*(*t*) cannot be explained by the assumed epidemic seed alone. Similarly, varying the immunity waning rate over a broad range alters the magnitude of the calibrated *κ*(*t*), particularly later in the epidemic, but preserves much of its overall temporal structure (Supplementary Section S2.3.4; Supplementary Figure S13). Together, these results suggest that *κ*(*t*) partly compensates for uncertainty in explicitly modelled processes, while persistent features across sensitivity analyses likely reflect residual dynamics not captured by those assumptions.

Some of these fluctuations coincide with major epidemiological and policy transitions. Early variability may reflect a combination of limited detected incidence, imported infections, incomplete representation of early behavioural change, and greater calibration uncertainty when fewer preceding weeks are available to the sliding window. Later changes in *κ*(*t*) occur around periods such as the return to school, variant emergence, and changes in reopening and travel policies. These temporal correspondences are consistent with changes in social mixing, reporting, and behavioural responses contributing to the inferred transmission multiplier, although *κ*(*t*) alone cannot attribute individual fluctuations to any one of these mechanisms. Future work could decompose residual variation in *κ*(*t*) by incorporating time-varying covariates that are currently implicit in calibration, such as mobility, testing intensity, variant prevalence, weather, and behavioural indicators. This could help distinguish identifiable external drivers from remaining unobserved dynamics and guide future model extensions.

The model reproduces overall seroprevalence trends but reveals challenges in mapping infections to antibodies. Anti-S closely matches observations, reflecting vaccination’s dominant contribution in 2021, whereas the anti-N comparison varies over time: the model underestimates the empirical estimate early in the study period and later lies above its lower uncertainty bound. This pattern may reflect imperfect mapping of infection to anti-N seropositivity, including seroconversion, waning, assay-threshold effects, and infection–vaccination-history heterogeneity. In addition, blood donors are a healthier, eligibility-screened group with higher vaccine uptake than the general population, and deferral policies exclude recent illness or exposure [59, 60]. Together, these factors can contribute to discrepancies between observed and model-based anti-N estimates. Similar model–data tensions are reported in national serosurveys and longitudinal cohorts [32, 61–64].

Several assumptions shape the interpretation of our counterfactuals. In our counterfactual analyses, we modify vaccination and NPIs, the latter through changes in the setting-specific contact matrices, while holding the calibrated *κ*(*t*) fixed. The calibrated *κ*(*t*) captures additional temporal variation in transmission, including seasonal and weather-related effects, changes in variant transmissibility, and behavioural changes such as waning compliance with protective measures. Many of these factors may evolve independently of the specific public health interventions imposed. For example, compliance with protective measures may naturally decline over time, even without changes in formal restrictions.

We recognize, however, that behavioural responses, government testing, contact tracing, and enforcement practices may also differ under alternative interventions. Determining how these factors would have evolved under each counterfactual scenario is difficult to establish from the available data. Holding *κ*(*t*) fixed therefore allows us to compare the effects of alternative public health interventions under common background transmission conditions without introducing additional poorly constrained assumptions. Reporting fractions for moderate/severe infections are assumed constant despite evolving testing and surveillance, and importations are omitted despite their growing role in late 2021. Although age-structured with setting-specific contacts and class-specific susceptibilities/infectivities, the model is effectively mean-field within age–immunity classes and spatially well-mixed; it omits explicit geography, household/workplace networks, clustering by vaccination or prior infection, quarantining, and superspreading. Accordingly, our results quantify the effects of alternative intervention strategies under common assumptions rather than predicting all behavioural and policy responses that might have occurred under each scenario.

Nonetheless, the study has notable strengths. While Fair et al. [28] use an age- and spatially structured stochastic model to examine NPIs in Ontario, our ODE framework tracks 16 age groups, vaccination status, and four immunity levels, allowing us to distinguish vaccine- from infection-derived protection and capture Ontario’s shift from naïve to vaccine-driven immunity. Calibration to both daily incidence and hospital admissions, complemented by an independent seroprevalence cross-check, provides a broader assessment of disease burden and population immunity than earlier Ontario studies focused on reported case incidence [21] or outbreaks in childcare centres and primary schools [27]. The ODE framework also enables efficient exploration of a broad range of counterfactuals.

Our counterfactual estimates are broadly consistent with previous Canadian modelling in showing that removing NPIs or vaccination substantially increases infections and hospital burden. However, a direct quantitative comparison with Ogden et al. [24], who use an agent-based model with 100,000 agents representing the Canadian population and incorporating vaccination and waning immunity, is limited. Our Ontario-specific model generally predicts larger cumulative infection totals, even though their analysis includes the Omicron wave whereas our comparison period does not. This difference likely reflects several structural and geographic differences, including Ontario-specific calibration, more population-level mixing in our deterministic framework, differences in household and venue structure, incidence-triggered isolation and quarantine mechanisms in the agent-based model, and different assumptions about symptom status and immunity transitions. The outcome-specific comparison further shows that the differences between the two models are more pronounced for asymptomatic and total infections than for hospital admissions (Supplementary Figure S19), suggesting that differences in infection classification and transmission structure may contribute to the discrepancies rather than a uniformly greater disease burden predicted by our model. In future work, we plan to extend our modelling framework to all of Canada, which should allow a more direct comparison with national-scale counterfactual analyses.

Beyond Ontario, this framework is portable. By parameterizing *C*_*n,m*_(*t*) with policy histories (e.g., OxCGRT stringency trajectories), we can test our counterfactuals against jurisdictions with contrasting NPI portfolios—such as Australia’s high-stringency, border-focused approach versus Sweden’s comparatively relaxed strategy—and evaluate how well predicted severe outcomes match observed experience [4]. The same machinery supports forward policy analysis for vaccination. For example, recent debates have considered limiting updated COVID-19 doses to older adults: the United States in August 2025 narrowed under-65 eligibility while maintaining full access for those ≥ 65 y, whereas the UK and EU programmes already target older or clinically vulnerable groups [65–67]. Setting age-targeted dosing schedules and appropriate initial conditions, the model can project hospital burden under alternative booster strategies (e.g., ≥60–65 y only vs. all adults) and identify thresholds where broader coverage materially reduces severe outcomes. These cross-jurisdictional and forward-looking applications also provide a basis for considering broader preparedness strategies for future respiratory pandemics.

For respiratory pandemics that disproportionately affect adults —as pre-Omicron COVID-19 did— preparedness may benefit from prioritizing measures that reduce contacts in high-density adult environments, alongside rapid vaccination and timely boosting of older or clinically vulnerable adults. Extended school closures may yield more limited epidemiological gains for adult-skewed pathogens while imposing substantial educational and social costs, and should therefore be used cautiously and selectively. Jurisdictions could pre-plan scalable, setting-specific NPIs linked to hospital or ICU indicators, while protecting high-risk settings through vaccination, outbreak control, paid sick leave, and clear risk communication. If pathogen severity shifts toward children, these priorities should be reconsidered.

## Acknowledgments

This work was supported by the Natural Sciences and Engineering Research Council of Canada (JMH) and the York University Research Chair Program (JMH).

## Data and Code Accessibility

The GitHub repository containing the code and data is available upon request.

## References

[1] Verity, R. et al. Estimates of the severity of coronavirus disease 2019: a model-based analysis. Lancet Infect. Dis. 20, 669–677 (2020).

[2] Nalbandian, A. et al. Post-acute COVID-19 syndrome. Nat. Med. 27, 601–615 (2021).

[3] World Health Organization. COVID-19 strategy update. Technical Report, World Health Organization, Geneva (2020). URL https://www.who.int/publications/i/item/covid-19-strategy-update14-april-2020. Accessed 2025-08-05.

[4] Hale, T. et al. A global panel database of pandemic policies (Oxford COVID-19 Government Response Tracker). Nat. Hum. Behav. 5, 529–538 (2021).

[5] Gebretekle, T. K. & Albers, C. Modelling the impact of behavioural interventions during pandemics: A systematic review. PLoS One 20, e0310611 (2025).

[6] Remuzzi, A. & Remuzzi, G. COVID-19 and Italy: what next? Lancet 395, 1225–1228 (2020).

[7] Hong, D., Yin, X. & Shi, L. The COVID catastrophe: what’s gone wrong and how to stop it happening again. Emerg. Infect. Dis. 27, 2976 (2021).

[8] Rothwell, J. T., Cojocaru, A., Srinivasan, R. & Kim, Y. S. Global evidence on the economic effects of disease suppression during COVID-19. Humanit. Soc. Sci. Commun. 11 (2024).

[9] Wisenoecker, A. S., Helm, C., Große, C. S., Hübner, N. & Zitzmann, S. A meta-analysis of students’ academic learning losses over the course of the COVID-19 pandemic. Learn. Instr. 98, 102111 (2025).

[10] Whitley, J., Beauchamp, M. H. & Brown, C. The impact of COVID-19 on the learning and achievement of vulnerable Canadian children and youth. Facets 6, 1693–1713 (2021).

[11] Abugamza, A., Kaskirbayeva, D., Charlwood, A., Nikolova, S. & Martin, A. Impact of the COVID-19 pandemic on employment and inequalities: a systematic review of international evidence and critical appraisal of statistical methods. Perspect. Public Health (2024).

[12] Jones, S. R., Lange, F., Craig Riddell, W. & Warman, C. The great Canadian recovery: the impact of COVID-19 on Canada’s labour market. Can. J. Econ. 56, 791–838 (2023).

[13] Mohseni, M. et al. An umbrella review of the prevalence of depression during the COVID-19 pandemic: call to action for post-COVID-19 at the global level. BMC Public Health 24, 3562 (2024).

[14] Cameron-Blake, E. et al. Variation in the stringency of COVID-19 public health measures on self-reported health, stress, and overall wellbeing in Canada. Sci. Rep. 13, 13094 (2023).

[15] He, X., Chen, H., Zhu, X. & Gao, W. Non-pharmaceutical interventions in containing COVID-19 pandemic after the roll-out of coronavirus vaccines: a systematic review. BMC Public Health 24, 1524 (2024).

[16] Brauner, J. M. et al. Inferring the effectiveness of government interventions against COVID-19. Science 371, eabd9338 (2021).

[17] Haug, N. et al. Ranking the effectiveness of worldwide COVID-19 government interventions. Nat. Hum. Behav. 4, 1303–1312 (2020).

[18] Mendez-Brito, A., El Bcheraoui, C. & Pozo-Martin, F. Systematic review of empirical studies comparing the effectiveness of non-pharmaceutical interventions against COVID-19. J. Infect. 83, 281–293 (2021).

[19] Liu, Y., Wang, W.Wong, W.-K. & Zhu, W. Effectiveness of non-pharmaceutical interventions for COVID-19 in USA. Sci. Rep. 14, 21387 (2024).

[20] Karaivanov, A., Lu, S. E., Shigeoka, H., Chen, C. & Pamplona, S. Face masks, public policies and slowing the spread of COVID-19: evidence from Canada. J. Health Econ. 78, 102475 (2021).

[21] Naimark, D. et al. Simulation-based estimation of SARS-CoV-2 infections associated with school closures and community-based nonpharmaceutical interventions in Ontario, Canada. JAMA Netw. Open 4, e213793–e213793 (2021).

[22] Marziano, V. et al. Retrospective analysis of the Italian exit strategy from COVID-19 lockdown. Proc. Natl. Acad. Sci. U.S.A. 118, e2019617118 (2021).

[23] Ogden, N. H. et al. Modelling scenarios of the epidemic of COVID-19 in Canada. Can. Commun. Dis. Rep. 46, 198–204 (2020).

[24] Ogden, N. H. et al. Counterfactuals of effects of vaccination and public health measures on COVID-19 cases in Canada: what could have happened? Can. Commun. Dis. Rep. 48, 292–302 (2022).

[25] Ludwig, A. et al. Assessing the impact of varying levels of case detection and contact tracing on COVID-19 transmission in Canada during lifting of restrictive closures using a dynamic compartmental model. Can. Commun. Dis. Rep. 46, 409–421 (2020).

[26] Hurford, A., Rahman, P. & Loredo-Osti, J. C. Modelling the impact of travel restrictions on COVID-19 cases in Newfoundland and Labrador. R. Soc. Open Sci. 8, 202266 (2021).

[27] Phillips, B., Browne, D. T., Anand, M. & Bauch, C. T. Model-based projections for COVID-19 outbreak size and student-days lost to closure in Ontario childcare centres and primary schools. Sci. Rep. 11, 6402 (2021).

[28] Fair, K. R., Karatayev, V. A., Anand, M. & Bauch, C. T. Estimating COVID-19 cases and deaths prevented by non-pharmaceutical interventions, and the impact of individual actions: a retrospective model-based analysis. Epidemics 39, 100557 (2022).

[29] Ng, V. et al. Modelling the impact of shutdowns on resurging SARS-CoV-2 transmission in Canada. R. Soc. Open Sci. 8, 210233 (2021).

[30] Shoukat, A. et al. Projecting demand for critical care beds during COVID-19 outbreaks in Canada. CMAJ 192, E489–E496 (2020).

[31] Iyaniwura, S. A. et al. Mathematical modeling of COVID-19 in British Columbia: an age-structured model with time-dependent contact rates. Epidemics 39, 100559 (2022).

[32] Dick, D. W. et al. COVID-19 seroprevalence in Canada modelling waning and boosting COVID-19 immunity in Canada a Canadian Immunization Research Network study. Vaccines 10, 17 (2022).

[33] Childs, L. et al. Modeling waning and boosting of COVID-19 in Canada with vaccination. Epidemics 39, 100583 (2022).

[34] Navazi, F., Yuan, Y. & Archer, N. The effect of the Ontario stay-at-home order on COVID-19 third wave infections including vaccination considerations: an interrupted time series analysis. PLoS One 17, e0265549 (2022).

[35] Akanteva, A., Dick, D. W., Amiraslani, S. & Heffernan, J. M. Canadian COVID-19 pandemic public health mitigation measures at the province level. Sci. Data 10, 882 (2023).

[36] Clark, A. et al. Global, regional, and national estimates of the population at increased risk of severe COVID-19 due to underlying health conditions in 2020: a modelling study. Lancet Glob. Health 8, e1003–e1017 (2020).

[37] Mohsin, M. & Mahmud, S. Omicron SARS-CoV-2 variant of concern: a review on its transmissibility, immune evasion, reinfection, and severity. Medicine 101, e29165 (2022).

[38] Government of Ontario. Ontario updating public health measures and guidance in response to Omicron (2022). URL https://news.ontario.ca/en/release/1001386/ontario-updating-public-health-measures-and-guidance-in-response-to-omicron. News release, accessed 2026-05-19.

[39] Berry, I., Soucy, J.-P. R., Tuite, A. & Fisman, D. Open access epidemiologic data and an interactive dashboard to monitor the COVID-19 outbreak in Canada. CMAJ 192, E420–E420 (2020).

[40] Public Health Agency of Canada. Canadian COVID-19 vaccination coverage report. https://health-infobase.canada.ca/covid-19/vaccination-coverage/ (2024). Accessed: 2026-07-06.

[41] Betti, M. I. et al. COVID-19 vaccination and healthcare demand. Bull. Math. Biol. 85, 32 (2023).

[42] Knight, J. & Mishra, S. Estimating effective reproduction number using generation time versus serial interval, with application to COVID-19 in the Greater Toronto Area, Canada. Infect. Dis. Model. 5, 889–896 (2020).

[43] Cleveland, W. S. & Devlin, S. J. Locally weighted regression: An approach to regression analysis by local fitting. J. Am. Stat. Assoc. 83, 596–610 (1988).

[44] Murphy, T. J. et al. The evolution of SARS-CoV-2 seroprevalence in Canada: a time-series study, 2020–2023. CMAJ 195, E1030–E1037 (2023).

[45] Verma, A. A. et al. Characteristics and outcomes of hospital admissions for COVID-19 and influenza in the Toronto area. CMAJ 193, E410–E418 (2021).

[46] Abdollahi, E., Haworth-Brockman, M., Keynan, Y., Langley, J. M. & Moghadas, S. M. Simulating the effect of school closure during COVID-19 outbreaks in Ontario, Canada. BMC Med. 18, 230 (2020).

[47] Kaffai, M. & Heiberger, R. H. Modeling non-pharmaceutical interventions in the COVID-19 pandemic with survey-based simulations. PLoS One 16, e0259108 (2021).

[48] El Jaouhari, M. et al. Impact of school closures and re-openings on COVID-19 transmission. Can. Commun. Dis. Rep. 47, 515–523 (2021).

[49] Levin, A. T. et al. Assessing the age specificity of infection fatality rates for COVID-19: systematic review, meta-analysis, and public policy implications. Eur. J. Epidemiol. 35, 1123–1138 (2020).

[50] O’Driscoll, M. et al. Age-specific mortality and immunity patterns of SARS-CoV-2. Nature 590, 140–145 (2021).

[51] UNESCO, UNICEF & World Bank. The state of the global education crisis: A path to recovery. Tech. Rep., UNESCO, UNICEF, and World Bank (2021).

[52] Racine, N. et al. Global prevalence of depressive and anxiety symptoms in children and adolescents during COVID-19: a meta-analysis. JAMA Pediatr. 175, 1142–1150 (2021).

[53] Ontario COVID-19 Science Advisory Table. School operation for the 2021–2022 academic year in the context of the COVID-19 pandemic. Science Briefs of the Ontario COVID-19 Science Advisory Table (2021).

[54] Walsh, S. et al. Do school closures and school reopenings affect community transmission of COVID-19? a systematic review of observational studies. BMJ Open 11, e053371 (2021).

[55] Monod, M. et al. Age groups that sustain resurging COVID-19 epidemics in the United States. Science 371, eabe8372 (2021).

[56] European Centre for Disease Prevention and Control. COVID-19 in children and the role of school settings in transmission – second update. Technical report, European Centre for Disease Prevention and Control, Stockholm (2021). URL https://www.ecdc.europa.eu/sites/default/files/documents/COVID-19-in-children-and-the-role-of-school-settings-in-transmission-second-update.pdf.

[57] Canadian Institute for Health Information. Children and youth with medical complexity in Canada. Tech. Rep., Canadian Institute for Health Information, Ottawa, ON (2020). URL https://www.cihi.ca/sites/default/files/document/children-youth-with-medical-complexity-report-en.pdf.

[58] The Hospital for Sick Children (SickKids). COVID-19: Guidance for school operation during the pandemic. Tech. Rep., The Hospital for Sick Children, Toronto, ON (2021). URL https://www.sickkids.ca/siteassets/news/news-archive/2021/covid19-guidance-for-school-operation-sickkids.pdf.

[59] Brodersen, T. et al. The healthy donor effect and survey participation, becoming a donor and donor career. Transfusion 63, 143–155 (2023).

[60] World Health Organization. Population-based age-stratified seroprevalence investigation template protocol for respiratory pathogens with pandemic potential. Template protocol, World Health Organization, Geneva (2023). URL https://cdn.who.int/media/docs/default-source/documents/epp/respiratory-unity-study/unity_studies-seroprevalence-investigation.pdf. Respiratory Investigations and Studies (Unity Studies); xLicence: CC BY-NC-SA 3.0 IGO.

[61] Molla, J., Farhang-Sardroodi, S., Moyles, I. R. & Heffernan, J. M. Pharmaceutical and nonpharmaceutical interventions for controlling the COVID-19 pandemic. R. Soc. Open Sci. 10, 230621 (2023).

[62] Van Elslande, J. et al. Lower persistence of anti-nucleocapsid compared to anti-spike antibodies up to one year after SARS-CoV-2 infection. Diagn. Microbiol. Infect. Dis. 103, 115659 (2022).

[63] Eyre, D. W. et al. Stringent thresholds in SARS-CoV-2 IgG assays lead to under-detection of mild infections. BMC Infect. Dis. 21, 187 (2021).

[64] Bobrovitz, N. et al. Global seroprevalence of SARS-CoV-2 antibodies: a systematic review and meta-analysis. PLoS One 16, e0252617 (2021).

[65] Reuters. US FDA narrows under-65 COVID vaccine eligibility; maintains full access for older Americans. News article (2025). URL https://www.reuters.com/business/healthcare-pharmaceuticals/us-fda-narrows-under-65-covid-vaccine-eligibility-maintains-full-access-older-2025-08-27/.

[66] Joint Committee on Vaccination and Immunisation. JCVI statement on the COVID-19 vaccination programme for autumn 2024: 8 April 2024. UK Government guidance (2024). URL https://www.gov.uk/government/publications/covid-19-autumn-2024-vaccination-programme-jcvi-advice-8-april-2024/jcvi-statement-on-the-covid-19-vaccination-programme-for-autumn-2024-8-april-2024.

[67] European Centre for Disease Prevention and Control. COVID-19 vaccination coverage in the EU/EEA during the 2024–25 season campaigns. Tech. Rep., ECDC, Stockholm (2025). URL https://www.ecdc.europa.eu/en/publications-data/covid-19-vaccination-coverage-eueea-during-2024-25-season-campaigns.

[68] Korosec, C. S. et al. Multiple cohort study of hospitalized SARS-CoV-2 in-host infection dynamics: parameter estimates, identifiability, sensitivity and the eclipse phase profile. J. Theor. Biol. 564, 111449 (2023).

[69] Movsisyan, M. et al. Tracking the evolution of anti-SARS-CoV-2 antibodies and long-term humoral immunity within 2 years after COVID-19 infection. Sci. Rep. 14, 13417 (2024).

[70] Shrotri, M. et al. Spike-antibody waning after second dose of BNT162b2 or ChAdOx1. Lancet 398, 385–387 (2021).

[71] Thomas, S. J. et al. Safety and efficacy of the BNT162b2 mRNA COVID-19 vaccine through 6 months. N. Engl. J. Med. 385, 1761–1773 (2021).

[72] Prem, K. et al. Projecting contact matrices in 177 geographical regions: an update and comparison with empirical data for the COVID-19 era. PLoS Comput. Biol. 17, e1009098 (2021).

[73] Public Health Ontario. COVID-19 real-world vaccine effectiveness: what we know so far. Tech. Rep., Queen’s Printer for Ontario, Toronto, ON (2021). URL https://www.publichealthontario.ca/-/media/documents/ncov/covid-wwksf/2021/04/wwksf-vaccine-effectiveness.pdf?la=en.

[74] Dan, J. M. et al. Immunological memory to SARS-CoV-2 assessed for up to 8 months after infection. Science 371, eabf4063 (2021).

[75] Davies, N. G. et al. Age-dependent effects in the transmission and control of COVID-19 epidemics. Nat. Med. 26, 1205–1211 (2020).

[76] Byambasuren, O. et al. Estimating the extent of asymptomatic COVID-19 and its potential for community transmission: systematic review and meta-analysis. J. Assoc. Med. Microbiol. Infect. Dis. Can. 5, 223–234 (2020).

[77] Sah, P. et al. Asymptomatic SARS-CoV-2 infection: a systematic review and meta-analysis. Proc. Natl. Acad. Sci. U.S.A. 118, e2109229118 (2021).

