## Supplementary Material for "Vaccination and Non-School NPIs Shaped Ontario’s Pre-Omicron COVID-19 Hospital Burden"

#### S1 Mathematical Model

We apply an extended Age-Structured Infection and Immunity Model (ASIIM) based on previous work [32, 33]. The earlier framework already incorporates age structure, multiple levels of infection- and vaccine-derived immunity, severity-dependent infection outcomes, waning immunity, vaccination, and age-specific contact patterns. Here, we extend this structure to explicitly track vaccination history within the susceptible, exposed, and infected populations, distinguishing individuals with and without prior vaccination ($S^V$, $E^V$, and $I^V$ versus $S$, $E$, and $I$), and to represent first-, second-, and booster-dose vaccination states separately ($V^1$, $V^2$, and $V^B$). This expanded structure allows vaccine eligibility, breakthrough infections, post-vaccination waning, and subsequent booster vaccination to be represented explicitly.

Age groups are indexed by $n = 1, 2, \ldots, N$, where $n = 1$ is the youngest and $n = N$ is the oldest. Figure S1 depicts the flow diagram for one age group. Individuals fall into one of four main compartments: *susceptible* ($S$), *exposed* ($E$), *infected* ($I$), and *vaccinated* ($V$). The $S$, $E$ and $I$ are further delineated into those with ($S^V$, $E^V$, $I^V$) and without ($S$, $E$, $I$) history of vaccination. Note that only $I$ and $I^V$ compartments generate new infections; $E$ and $E^V$ compartments are infected but not yet infectious.

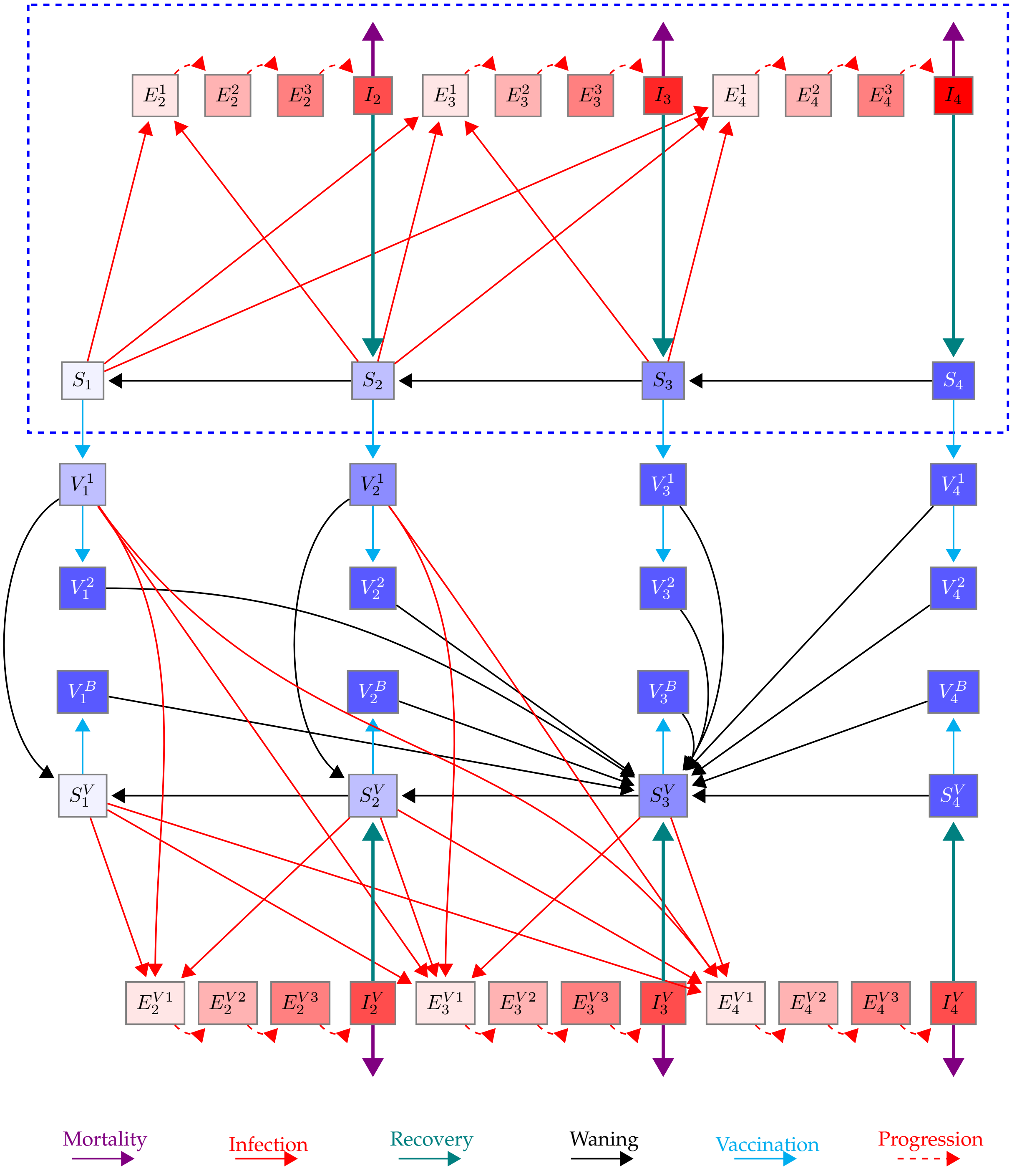


Figure S1: Model flow diagram for one age group, showing infection, progression, recovery, vaccination, waning, and mortality transitions among susceptible, infected, vaccinated, and immune compartments.

#### S1.1 Description of State Variables

The model state variables are indexed by age group ($n$) and either (1) immunity status ($i = 1$–$4$, from fully susceptible to fully resistant) or (2) infection severity ($j = 2$–$4$, mild to severe). For susceptible, exposed, and infected classes, the superscript $V$ indicates prior vaccination (e.g., $S_i^V$ are level-$i$ susceptibles with vaccination history where , $i = 1, \ldots, 4$ denotes immunity status). The distinction between compartments with prior vaccination and those without vaccination history allows for a more accurate determination of vaccine eligibility. Exposed individuals progress through three stages ($E^1$, $E^2$, $E^3$) before becoming infectious ($I$). Vaccinated compartments also carry a superscript denoting dose (1 = first, 2 = second, $B$ = booster).

Table S1 defines all state variables in the model and clarifies the indexing scheme. We note that we use $S_i = \sum_{n=1}^{N} S_{i,n}$ to denote all unvaccinated susceptibles in immunity class $i$. Likewise, omitting the immunity-level index $i$ or the severity index $j$ refers to all corresponding classes; for instance, $I^V = \sum_{n=1}^{N} \sum_{j=2}^{4} I_{j,n}^V$ represents all vaccinated infections over all ages combined.

| Compartment | Definition |
|---|---|
| $S_{i,n}$ | Susceptible, age group $n$, immunity level $i$ |
| $S_{i,n}^V$ | Vaccinated Susceptible (post-vaccine waning), age group $n$, immunity level $i$ |
| $I_{j,n}$ | Infected, age $n$, symptom severity $j$ |
| $I_{j,n}^V$ | Vaccinated infected, age $n$, symptom severity $j$ |
| $E_{j,n}^k$ | Exposed stage $k$, destined for $I_{j,n}$ |
| $E_{j,n}^{V\,k}$ | Vaccinated exposed stage $k$, destined for $I_{j,n}^V$ |
| $V_{i,n}^1$ | Vaccinated with one dose, age $n$, immunity level $i$ |
| $V_{i,n}^2$ | Fully vaccinated (two doses), age $n$, immunity level $i$ |
| $V_{i,n}^B$ | Vaccinated with booster, age $n$, immunity level $i$ |

Table S1: Model compartments and indexing notation. Here, $n = 1, \ldots, N$ denotes age group, $i = 1, \ldots, 4$ denotes immunity level, $j = 2, 3, 4$ denotes symptom severity class, and $k = 1, 2, 3$ denotes exposed-stage progression.

#### S1.2 Overall Flow

It is assumed that the population is fully susceptible at time zero and that there is a small infection seed. Unvaccinated susceptibles, $S_{i,n}$, can either become infected ($E_{j,n}^1 \to E_{j,n}^2 \to E_{j,n}^3 \to I_{j,n}$, with $j = 2, 3, 4$) or transition into the first-dose vaccination compartments, $V_{i,n}^1$. We assume that (1) the severity of infection, $j$, is determined by comorbidity-based probabilities [36], and that (2) the level of infection-induced immunity corresponds to the infection severity [68, 69]. In other words, individuals without comorbidities develop mild infections and recover with some immunity (level 2), those with exactly one comorbidity develop moderate infections and get moderate immunity upon recovery (level 3), and those with two or more comorbidities develop severe infections and then become fully resistant if they survive (level 4). It is assumed that severe infections will definitely show symptoms, but that mild and moderate infections show no or some mild symptoms, respectively. When individuals have recovered from infection, they will experience waning immunity. Immunity wanes between susceptible compartments from higher to lower levels, depending on immunity class and age group.

After receiving the first dose, individuals are not yet fully protected from infection unless they already have moderate immunity ($i = 3$) or are fully resistant ($i = 4$). For first-dose vaccinated individuals, immunity can wane to the corresponding vaccinated susceptible compartments at the same immunity level, except for those that are fully resistant, $V_4^1$, that wane to $S_3^V$. If individuals in $V^1$ neither become infected nor wane before receiving the second dose, they gain full protection through vaccination and move to $V^2$.

Vaccinated susceptibles, $S^V_{i,n}$, can become infected ($E^{V1}_{j,n} \rightarrow E^{V2}_{j,n} \rightarrow E^{V3}_{j,n} \rightarrow I^V_{j,n}$, with $j = 2, 3, 4$) or move into the booster compartment, $V^B_{i,n}$. Second- and booster-dose vaccinated individuals are assumed to be fully protected from infection (similar to $S^V_4$) and therefore always wane to $S^V_3$.

#### S1.3 Model Parameters

Table S2 lists the model parameters, their values, and their descriptions. Most parameter values are justified in previous work by our group [32, 33]; here, we build on that work.

##### S1.3.1 Disease Parameters

Disease progression in the model depends on immunity status, symptom severity, and age. Susceptibility, $\alpha_{i,n}$, decreases with increasing immunity. Infectivity, $\beta_{j,n}$, varies with disease severity and is scaled to produce a basic reproduction number of $R_0 = 2.6$ [42], with milder cases transmitting less due to lower viral loads and more severe cases transmitting less due to reduced mobility as it is assumed that they stay home from work or are admitted to healthcare. Recovery rates, $\gamma_{j,n}$, also depend on severity, with milder cases recovering more quickly and severe cases experiencing longer infectious periods. [68, 74].

The probability of progressing to more severe disease increases with age, which serves as a proxy for comorbidities. We use comorbidity data from Clark et al. [36] to determine the probability $p^j_{i,n}$ of developing a disease of severity $j$, given immunity level $i$ and age group $n$. For Canada, the values are provided in Table S3 and are applied for $i = 1$ and $i = 2$. For $i = 3$, it is assumed that movement to infection status $i = 2$ is infeasible as individuals that recover would recover from infection with less immunity than they started with. Instead, we assume that $i = 3$ individuals can have such a mild infection that they do not transmit infection and their immunity status does not change i.e., they remain in $S_3$. For $i = 4$, all probabilities are set to zero, since individuals with immunity level 4 are fully protected against infection.

Once individuals are partially vaccinated, i.e., in $V^1$, they can still become infected if their immunity level is $i = 1$ or $i = 2$, but with protection of $1 - q$ against severe disease. The adjusted probabilities are given in Table S4. If the immunity of $V^1$ individuals wanes before they receive the second dose, they go to the vaccinated susceptible class, and the severity probabilities revert to $p^j_{i,n}$ (Table S3).

##### S1.3.2 Contact Rates and Force of Infection

The age-specific contact rates that underlie transmission in the model are derived from a pre-pandemic Ontario contact matrix, stratified into 16 age groups. The force of infection for age group $n$ at time $t$ is defined as

$$\lambda_n(t) \ = \ \kappa(t) \sum_{m=1}^{N} c_{n,m}(t) \sum_{j=2}^{4} \beta_{j,m} \frac{I_{j,m}(t) + I^V_{j,m}(t)}{N_m},$$

where $\kappa(t)$ is a time-varying scaling parameter calibrated to observed incidence and/or hospitalization data, $c_{n,m}(t)$ is the fraction of contacts between age groups $n$ and $m$, $I_{j,m}(t)$ and $I^V_{j,m}(t)$ are the numbers of unvaccinated and vaccinated infectious individuals of severity class $j$ in age group $m$, $N_m$ is the total population in age group $m$, and $\beta_{j,m}$ is the relative infectivity of severity class $j$ for age group $m$.

To calculate the contact fraction matrix $c_{n,m}$ and the total contacts $A_n$, we follow a multi-step procedure. First, each baseline contact matrix is converted from the per-capita rates -provided by [72]- to absolute contacts by multiplying each row $n$ by the corresponding population $N_n$, then symmetrized to ensure reciprocity (so that contacts reported between groups $n$ and $m$ are consistent in both directions), and finally returned to a per-capita basis.

| Parameter | Definition | Value/Comment | Source |
|---|---|---|---|
| $\alpha_{i,n}$ | Susceptibility of immunity class $i$ in age group $n$ | $\alpha_{1,n} = 1,\ \alpha_{2,n} = \frac{2}{3},\ \alpha_{3,n} = \frac{1}{3}$ for all $n$ | [33] |
| $\beta_{j,n}$ | Infectivity of severity class $j$ in age group $n$ | $\beta_{2,n} = 0.5\beta_{3,n},\ \beta_{3,n} = 0.0545,\ \beta_{4,n} = 0.1\beta_{3,n}$ for all $n$<br>Calibrated to $R_0 = 2.6$ | [42] |
| $\gamma_{j,n}$ | Recovery rate of severity class $j$ in age group $n$ | $\gamma_{2,n} = \frac{1}{5},\ \gamma_{3,n} = \frac{1}{10},\ \gamma_{4,n} = \frac{1}{15}$ for all $n$ | [33] |
| $\omega_{i,n}$ | Waning rate of immunity class $i$ in age group $n$ | $\omega_{i,n} = \frac{1}{365}$ for all $i$ and $n$ | [33], [70], [71] |
| $A_n$ | Per capita contact rate within age group $n$ | Calculated from contact matrices | [72] |
| $c_{n,m}$ | Fraction of all contacts of an individual in group $n$ that occur with individuals in group $m$ | Calculated from contact matrices | [72] |
| $\zeta_{j,n}$ | Progression rate between exposed classes in severity $j$ and age group $n$ | $\zeta_{j,n} = \frac{2}{3}$ for all $j$ and $n$ | [33] |
| $\sigma^{\ell}_{i,n}$ | Vaccination rate to dose $\ell$ for class $i, n$ | Based on vaccine coverage data | |
| $\epsilon$ | Susceptibility reduction due to partial vaccination | 30% | conservative estimate from [73] |
| $q$ | Residual fraction of severe-disease risk after partial vaccination | 0.08 | [33] |
| $\rho$ | Dose take probability | 0.9 | [32] |
| $p^{j}_{i,n}$ | Probability of severity $j$ given infection from class $i, n$ | Table S3<br>Based on comorbidities | [36] |
| $\bar{p}^{j}_{i,n}$ | Probability of severity $j$ in partially vaccinated individuals of class $i, n$ | Table S4 | [36], [33] |
| $\tau_n$ | aging rate for group $n$ | $\tau_n = \frac{1}{(5)(365)}$<br>(age groups are 5-year intervals) | |
| $\mu_n$ | Natural mortality rate in age group $n$ | 0<br>Ignored | |
| $\delta_{j,n}$ | Disease-induced mortality rate of severity $j$ in group $n$ | 0<br>Ignored | |
| $\eta$ | Birth rate | 0<br>Ignored | |
| $I_0$ | Initial condition for the infectious classes in the selected 6 age groups | $I_0 = \frac{1}{6}$ | |

Table S2: Model parameters, definitions, values, and sources used in the Ontario calibration and counterfactual simulations.

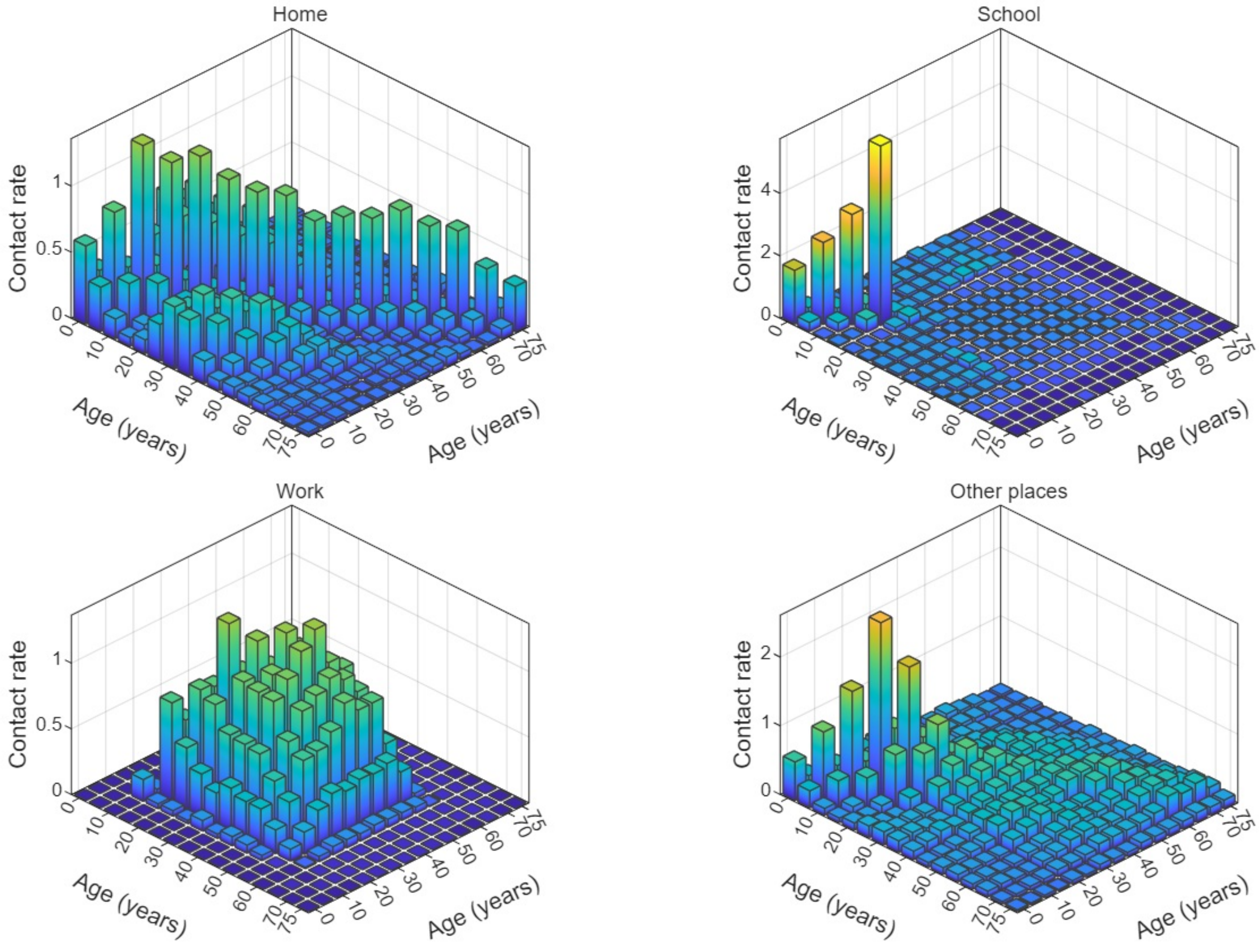


Figure S2: Baseline age-structured contact matrices for home, school, workplace, and other settings in the absence of NPIs. Bar heights represent contact rates between age groups.

| Group ($n$) | Age Range | $p^2_{i,n}$ | $p^3_{i,n}$ | $p^4_{i,n}$ |
|---|---|---|---|---|
| 1 | 0–4 | 0.979187625 | 0.019532353 | 0.001280022 |
| 2 | 5–9 | 0.970340674 | 0.027954180 | 0.001705146 |
| 3 | 10–14 | 0.971354725 | 0.026962603 | 0.001682673 |
| 4 | 15–19 | 0.963790465 | 0.033836424 | 0.002373111 |
| 5 | 20–24 | 0.947364080 | 0.048618385 | 0.004017534 |
| 6 | 25–29 | 0.923743993 | 0.069289774 | 0.006966234 |
| 7 | 30–34 | 0.897203802 | 0.091513710 | 0.011282488 |
| 8 | 35–39 | 0.865605311 | 0.116760519 | 0.017634170 |
| 9 | 40–44 | 0.806984827 | 0.162542801 | 0.030472373 |
| 10 | 45–49 | 0.756587998 | 0.197861718 | 0.045550284 |
| 11 | 50–54 | 0.690245134 | 0.241501371 | 0.068253495 |
| 12 | 55–59 | 0.600190415 | 0.296345592 | 0.103463993 |
| 13 | 60–64 | 0.503245046 | 0.346804634 | 0.149950320 |
| 14 | 65–69 | 0.409505408 | 0.383960071 | 0.206534521 |
| 15 | 70–74 | 0.324664092 | 0.404030163 | 0.271305745 |
| 16 | 75+ | 0.215150255 | 0.373779207 | 0.411070537 |

Table S3: Age-specific probabilities of mild, moderate, and severe infection, denoted by $p^2_{i,n}$, $p^3_{i,n}$, and $p^4_{i,n}$, respectively. These probabilities are based on age-specific comorbidity profiles from [36] and are applied directly at immunity levels $i = 1, 2$. The level-3 and level-4 exceptions are specified in the model assumptions.

| | $j = 2$ | $j = 3$ | $j = 4$ |
|---|---|---|---|
| $\bar{p}^j_{2,n}$ | $p^1_{2,n} + \left(1 - \frac{1}{2}q\right) p^2_{2,n}$ | $(1-q)p^3_{2,n} + \frac{1}{2}qp^2_{2,n}$ | $qp^3_{2,n}$ |
| $\bar{p}^j_{3,n}$ | 0 | $(1-q)p^3_{3,n} + \frac{1}{2}qp^2_{3,n}$ | $qp^3_{3,n}$ |

Table S4: Vaccine-adjusted severity probabilities for partially vaccinated individuals. The adjusted probabilities $\bar{p}^j_{i,n}$ incorporate the residual severe-disease fraction $q$ after partial vaccination; equivalently, $1 - q$ represents protection against severe disease.

The resulting matrices (shown in figure S2) are then modified by setting-specific perturbation functions that capture public health closures at each phase (school, work, other):

$$C_{n,m}(t) \;=\; C^{\text{home}}_{n,m} + \phi_{\text{school}}(t)\, C^{\text{school}}_{n,m} + \phi_{\text{work}}(t)\, C^{\text{work}}_{n,m} + \phi_{\text{other}}(t)\, C^{\text{other}}_{n,m}.$$

Here, $C^{\text{home}}_{n,m}$, $C^{\text{school}}_{n,m}$, $C^{\text{work}}_{n,m}$, and $C^{\text{other}}_{n,m}$ represent the baseline matrices for each setting, while $\phi_{\text{school}}(t)$, $\phi_{\text{work}}(t)$, and $\phi_{\text{other}}(t)$ are discrete scaling functions that perturb the matrices according to the public health measures in place. These are age-specific percentages based on the labels $\{0, 1, 2, 3\}$, corresponding to no restrictions (0), mild restrictions (1), moderate restrictions (2), and maximum closure (3). The time series of these closure levels is obtained from Akanteva et al. [35]. The corresponding perturbation matrices used for each closure level are provided in figure S3. Household contacts are assumed constant over time.

From the scaled contact matrix, the total number of contacts made by age group $n$ is

$$A_n = \sum_{m=1}^{N} C_{n,m},$$

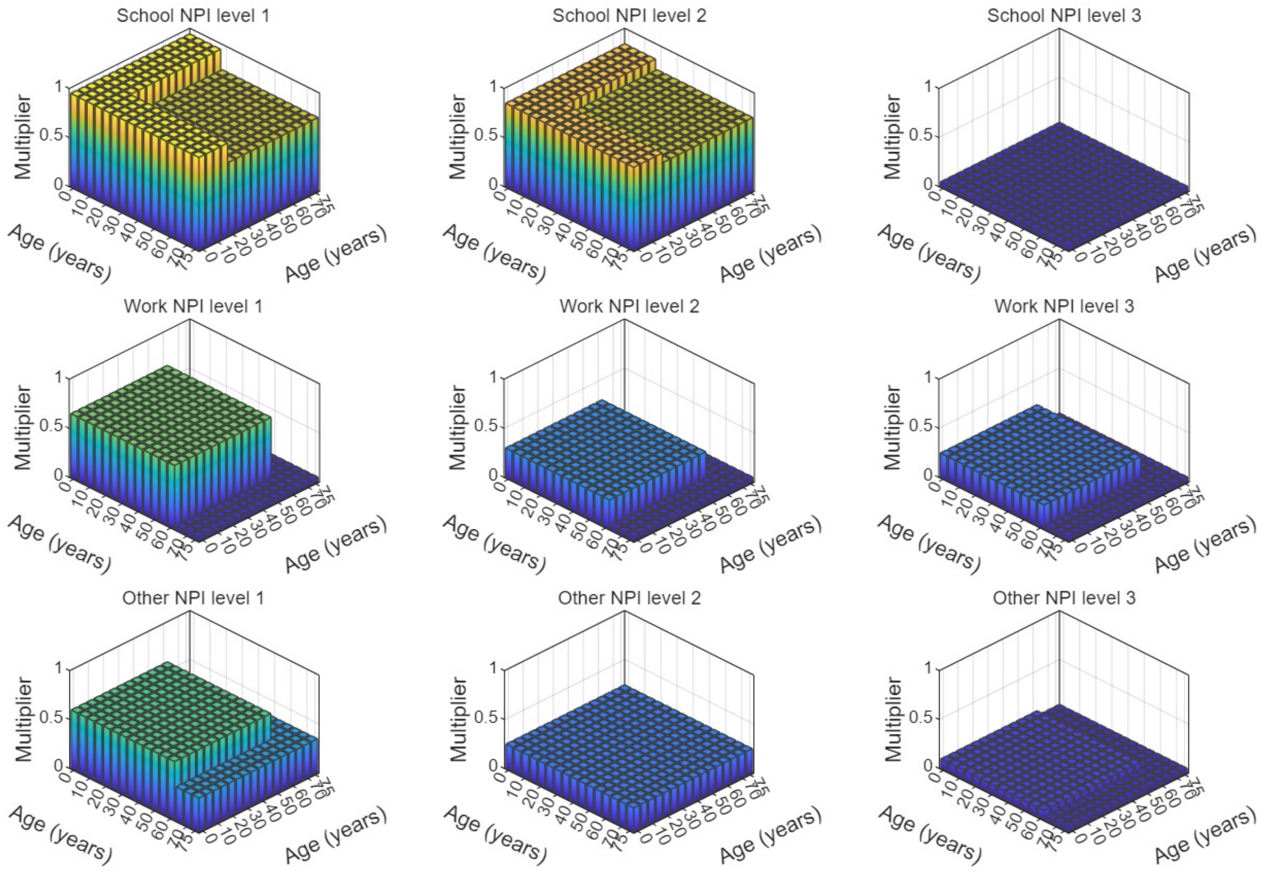


Figure S3: Contact perturbation matrices used to represent NPI levels in school, workplace, and other settings. Values are multipliers applied to the corresponding baseline contact matrices.

and the fraction of those contacts directed to group $m$ is

$$c_{n,m} = \frac{C_{n,m}}{A_n}, \qquad \sum_{m=1}^{N} c_{n,m} = 1.$$

Thus, $A_n$ quantifies overall contact activity for group $n$, while $c_{n,m}$ specifies how those contacts are distributed across other groups.

##### *S1.3.3 Vaccination Parameters*

We construct age- and dose-specific vaccination rates by translating aggregate coverage targets and policy-defined eligibility into per-step probabilities and then continuous-time rates over the mitigation window. The procedure uses two data streams: (i) overall (non–age-resolved) vaccine coverage, which provides the cumulative targets to match, and (ii) vaccine policy schedules that specify which age groups are eligible at each time; when eligibility is limited to health-care workers, we operationalize this by allowing vaccination of ages 20+ (because the model lacks occupation strata), but only for a small fraction of this population approximately given by (# of healthcare workers) / (total 20+ population).

For each age and dose, we compute the remaining gap to the target and scale it by the size of the dose-appropriate eligible pool in that age: for first doses, eligible unvaccinated susceptible individuals; for second doses, individuals who have received exactly one prior dose; for boosters, vaccinated-susceptible classes. The resulting vaccination probability over each model time step is then converted into a continuous-time vaccination rate, so that vaccination is applied smoothly over time rather than as a single discrete jump at each step. These rates are applied uniformly across the four immunity levels within each age, and rescaled to respect a campaign-level capacity constraint.

In the model, we distinguish three parameters related to vaccine effects. The parameter $\rho$ is the *dose take probability*—the probability that an administered dose successfully moves an individual into the corresponding vaccinated class because they acquire immunity from the vaccine; we apply $\rho$ to first and booster doses only, while second doses are assumed to take with certainty once the first dose has been received. The parameter $\epsilon$ encodes the *additional reduction in susceptibility for partially vaccinated individuals* (first dose), scaling their infection hazard relative to unvaccinated susceptibles. Finally, $q$ represents the residual fraction of severe-disease risk for partially vaccinated individuals. Thus, $1 - q$ represents protection against severe disease. This adjustment is applied only to infections that occur while individuals are in the first-dose vaccination compartments, $V^1$. It is not applied to infections occurring after individuals have waned into the vaccinated-susceptible compartments, $S^V$; for those infections, the baseline age- and immunity-specific severity probabilities are used.

##### *S1.3.4 Demographics*

We allow the model to accommodate demographic changes such as aging, births, and deaths, including both natural and disease-induced mortality. However, for the purpose of this work, which simulates only the first two years of the pandemic, we do not include any demographic changes other than aging. Disease-induced mortality is set to zero, since deaths represent a very small proportion of the total population and are not part of the calibration targets. Excluding mortality ensures that individuals progress through infection to recovery and into post-infection immunity classes without being removed from the system, thereby simplifying the compartment structure while preserving the dynamics relevant to incidence, hospitalization, and immunity. Aging is retained because it directly affects vaccine eligibility across age groups.

Moreover, for each immunity class $i$ or severity class $j$, the model can assign age-specific values for susceptibility ($\alpha_{i,n}$), infectivity ($\beta_{j,n}$), recovery rate ($\gamma_{j,n}$), progression rate ($\zeta_{j,n}$), and immunity waning rate ($\omega_{i,n}$). For the purpose of this work, however, we assume these parameters are uniform across age groups.

##### *S1.3.5 Initial Conditions*

We initialize the fully susceptible population ($S_1$) across age groups using Ontario's demographic distribution. A single imported case is introduced by seeding the infectious classes ($I$), but we distribute this seed across age groups. Specifically, we consider six candidate age groups—the 5th through 10th of 16 total (i.e., ages 20–50)—and assign each a probability of $I_0 = 1/6$ for the first infection. Within any selected age group, $I_0$ is then apportioned according to that group's three severity probabilities. Correspondingly, to preserve the total population size, we subtract $I_0$ from the susceptibles in each of these age groups. A sensitivity analysis of the initial condition is presented in Section S2.3.3.

##### *S1.3.6 Waning Rate*

Building on prior work [32, 33], we adopt a one-year waning rate from each higher immunity class to the next lower. This corresponds to $\omega = 1$ year$^{-1}$, or an average waning time of $T = 1$ year. This rate is assumed to be identical across age groups, and although waning could vary by vaccination or susceptibility status, we use a single common waning rate for unvaccinated susceptibles, vaccinated susceptibles, and vaccinated compartments. Sensitivity analysis to the waning rate is shown in section S2.3.4.

##### *S1.3.7 Calibration Proportions*

To align model outputs with observed incidence and hospitalization data, we derive daily incidence from the flow of individuals progressing from the final exposed stage into the corresponding infectious compartments. We assume that all incident severe infections are detected, 60% of incident moderate infections are detected, and mild infections are not systematically detected. Daily hospital admissions are estimated as 15% of daily severe incidence. These proportions are applied consistently across age groups. A sensitivity analysis of these calibration parameters is presented in Section S2.3.1.

#### *S1.4 Seroprevalence Estimation*

Model-predicted immunity distributions can be compared qualitatively with Canadian seroprevalence estimates. Seroprevalence measures antibody positivity, whereas the model's immunity distributions represent broader protection that may include cell-mediated and innate immune components. Anti-nucleocapsid (anti-N) antibodies indicate prior infection, whereas anti-spike (anti-S) antibodies indicate prior infection or vaccination [44]. Accordingly, Anti-S is compared against all susceptible compartments with immunity reflecting prior spike exposure: unvaccinated recovered layers $S_2 + S_3 + S_4$, vaccinated susceptibles $\sum_{i=1}^{4} S_i^V$, and the transient vaccinated compartments $\sum_{i=1}^{4}(V_i^1 + V_i^2 + V_i^B)$.

Anti-N is more complicated, since the sum of the recovered susceptibles $S_i, S_i^V,\ i = 2, 3, 4$ is confounded by vaccination history, as waning from fully vaccinated and booster compartments feeds into $S_3^V$. Thus, instead of providing a direct estimate of anti-N, we report an upper bound given by $\sum_{i=2}^{4} S_i + S_i^V$ and a lower bound given by the unvaccinated recovered classes only, $\sum_{i=2}^{4} S_i$. The true anti-N estimate may then lie between these bounds, including all unvaccinated recovered classes $\sum_{i=2}^{4} S_i$ and only a fraction of the vaccinated recovered classes, $\sum_{i=2}^{4} S_i^V$. Again, we note however that the model immunity distributions do not represent only antibody-induced immunity and thus they should not be expected to agree entirely.

For each age group, we sum the specified compartments and normalize by the age-group population to obtain model-based comparisons of anti-N and anti-S trajectories that are consistent with the empirical serology definitions.

To ensure comparability with Canadian Blood Services data, which are derived from blood donors aged 17 years and older, we present model seroprevalence results only for ages 15 and above, as our model divides the population into 5-year age intervals.

#### S1.5 Model Equations

In the following ODEs, indices are $i = 1, 2, 3, 4$ (immunity), $j = 2, 3, 4$ (symptom severity), $k = 1, 2, 3$ (exposed stage), $n = 1, \ldots, N$ (age group), $l = 1, 2, B$ (vaccine dose). To maintain consistency in the equations, we assume $\tau_0 = 0$ and that any compartment indexed with $n = 0$ contains zero individuals, even though no age group is actually indexed as zero. Similarly, since the oldest age group does not age further, we set $\tau_N = 0$. Let

$$\Lambda_{i,n}(t) = \alpha_{i,n} A_n \lambda_n(t),$$
$$\bar{\Lambda}_{i,n}(t) = (1-\epsilon)\alpha_{i,n} A_n \lambda_n(t).$$

*Unvaccinated Susceptible.*

$$\begin{aligned}
\frac{dS_{1,n}}{dt} =& \tau_{n-1}S_{1,n-1} - (\tau_n + \mu_n + \sigma^1_{1,n}\rho)S_{1,n} - \sum_{j=2}^{4} p^j_{1,n}\Lambda_{1,n}S_{1,n} + \omega_{2,n}S_{2,n}, \\
\frac{dS_{2,n}}{dt} =& \tau_{n-1}S_{2,n-1} - (\tau_n + \mu_n + \omega_{2,n} + \sigma^1_{2,n}\rho)S_{2,n} - \sum_{j=2}^{4} p^j_{2,n}\Lambda_{2,n}S_{2,n} + \omega_{3,n}S_{3,n} + \gamma_{2,n}I_{2,n}, \\
\frac{dS_{3,n}}{dt} =& \tau_{n-1}S_{3,n-1} - (\tau_n + \mu_n + \omega_{3,n} + \sigma^1_{3,n}\rho)S_{3,n} - \sum_{j=2}^{4} p^j_{3,n}\Lambda_{3,n}S_{3,n} + \omega_{4,n}S_{4,n} + \gamma_{3,n}I_{3,n}, \\
\frac{dS_{4,n}}{dt} =& \tau_{n-1}S_{4,n-1} - (\tau_n + \mu_n + \omega_{4,n} + \sigma^1_{4,n}\rho)S_{4,n} + \gamma_{4,n}I_{4,n},
\end{aligned}$$

*Vaccinated Susceptible.*

$$\begin{aligned}
\frac{dS^V_{1,n}}{dt} =& \tau_{n-1}S^V_{1,n-1} - (\tau_n + \mu_n + \sigma^B_{1,n}\rho)S^V_{1,n} - \sum_{j=2}^{4} p^j_{1,n}\Lambda_{1,n}S^V_{1,n} + \omega_{2,n}S^V_{2,n} + \omega_{1,n}V^1_{1,n}, \\
\frac{dS^V_{2,n}}{dt} =& \tau_{n-1}S^V_{2,n-1} - (\tau_n + \mu_n + \omega_{2,n} + \sigma^B_{2,n}\rho)S^V_{2,n} - \sum_{j=2}^{4} p^j_{2,n}\Lambda_{2,n}S^V_{2,n} + \gamma_{2,n}\, I^V_{2,n} + \omega_{3,n}S^V_{3,n} + \omega_{2,n}V^1_{2,n}, \\
\frac{dS^V_{3,n}}{dt} =& \tau_{n-1}S^V_{3,n-1} - (\tau_n + \mu_n + \omega_{3,n} + \sigma^B_{3,n}\rho)S^V_{3,n} - \sum_{j=2}^{4} p^j_{3,n}\Lambda_{3,n}S^V_{3,n} + \gamma_{3,n}\, I^V_{3,n} + \omega_{4,n}S^V_{4,n} \\
&+ \omega_{3,n}\Big(V^1_{3,n} + V^1_{4,n} + V^2_{1,n} + V^2_{2,n} + V^2_{3,n} + V^2_{4,n} + V^B_{1,n} + V^B_{2,n} + V^B_{3,n} + V^B_{4,n}\Big), \\
\frac{dS^V_{4,n}}{dt} =& \tau_{n-1}S^V_{4,n-1} - (\tau_n + \mu_n + \omega_{4,n} + \sigma^B_{4,n}\rho)S^V_{4,n} + \gamma_{4,n}\, I^V_{4,n}.
\end{aligned}$$

*Exposed and vaccinated exposed.*

$$\frac{\mathrm{d}E_{j,n}^{k}}{\mathrm{d}t} = \begin{cases} \tau_{n-1}E_{j,n-1}^{k} - (\tau_n + \mu_n + \zeta_{j,n})E_{j,n}^{k} + \sum_{i=1}^{3} p_{i,n}^{j}\Lambda_{i,n}S_{i,n}, & k = 1 \\ \tau_{n-1}E_{j,n-1}^{k} - (\tau_n + \mu_n)E_{j,n}^{k} + \zeta_{j,n}(E_{j,n}^{k-1} - E_{j,n}^{k}), & k = 2,3 \end{cases}$$

$$\frac{\mathrm{d}E_{j,n}^{V\,k}}{\mathrm{d}t} = \begin{cases} \tau_{n-1}E_{j,n-1}^{V\,k} - (\tau_n + \mu_n + \zeta_{j,n})E_{j,n}^{V\,k} + \sum_{i=1}^{3} p_{i,n}^{j}\Lambda_{i,n}S_{i,n}^{V} \\ \qquad + \sum_{i=1}^{2} \bar{p}_{i,n}^{j}\bar{\Lambda}_{i+1,n}V_{i,n}^{1}, & k = 1 \\ \tau_{n-1}E_{j,n-1}^{V\,k} - (\tau_n + \mu_n)E_{j,n}^{V\,k} + \zeta_{j,n}(E_{j,n}^{V\,k-1} - E_{j,n}^{V\,k}), & k = 2,3 \end{cases}$$

*Infected and vaccinated infected.*

$$\frac{\mathrm{d}I_{j,n}}{\mathrm{d}t} = \tau_{n-1}I_{j,n-1} - (\tau_n + \mu_n + \delta_{j,n} + \gamma_{j,n})I_{j,n} + \zeta_{j,n}E_{j,n}^{3}$$

$$\frac{\mathrm{d}I_{j,n}^{V}}{\mathrm{d}t} = \tau_{n-1}I_{j,n-1}^{V} - (\tau_n + \mu_n + \delta_{j,n} + \gamma_{j,n})I_{j,n}^{V} + \zeta_{j,n}E_{j,n}^{V\,3}$$

*Vaccinated – first, second and booster doses.*

$$\frac{\mathrm{d}V_{i,n}^{1}}{\mathrm{d}t} = \begin{cases} \tau_{n-1}V_{i,n-1}^{1} - (\tau_n + \mu_n + \omega_{i,n} + \sigma_{i,n}^{2})V_{i,n}^{1} + \sigma_{i,n}^{1}\rho S_{i,n} \\ \qquad - \sum_{j=2}^{4} \bar{p}_{i+1,n}^{j}\bar{\Lambda}_{i+1,n}V_{i,n}^{1}, & i = 1,2 \\ \tau_{n-1}V_{i,n-1}^{1} - (\tau_n + \mu_n + \omega_{i,n} + \sigma_{i,n}^{2})V_{i,n}^{1} + \sigma_{i,n}^{1}\rho S_{i,n}, & i = 3,4 \end{cases}$$

$$\frac{\mathrm{d}V_{i,n}^{2}}{\mathrm{d}t} = \tau_{n-1}V_{i,n-1}^{2} - (\tau_n + \mu_n + \omega_{i,n})V_{i,n}^{2} + \sigma_{i,n}^{2}V_{i,n}^{1}$$

$$\frac{\mathrm{d}V_{i,n}^{B}}{\mathrm{d}t} = \tau_{n-1}V_{i,n-1}^{B} - (\tau_n + \mu_n + \omega_{i,n})V_{i,n}^{B} + \sigma_{i,n}^{B}\rho S_{i,n}^{V}$$

### S2 Supplementary Results

#### *S2.1 Calibration under alternative data targets*

We first examine how the model calibration changes when incidence and hospital-admission data are calibrated separately rather than jointly. When the model is calibrated to incidence alone, the observed incidence trajectory is closely reproduced, whereas larger discrepancies remain in hospital admissions, particularly during the first epidemic wave (Figure S4). Conversely, calibration to hospital admissions alone reproduces the hospitalization trajectory more closely but results in larger discrepancies in incidence, most notably during the first wave (Figure S5).

The corresponding residuals illustrate this trade-off (Figure S6). As expected, calibrating to a single data stream produces the smallest residual for that particular target. However, simultaneous calibration to incidence and hospital admissions provides the lowest combined residual and a more balanced calibration across both outcomes. We therefore use the joint calibration for the counterfactual analyses presented in the main manuscript.

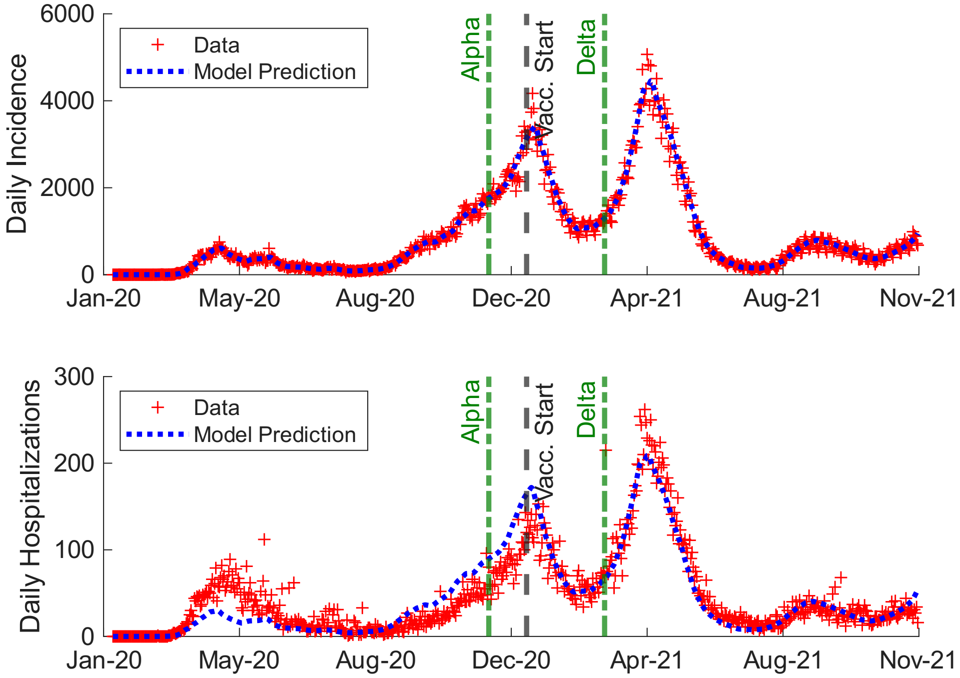


Figure S4: Model calibrated to incidence data only. Both incidence (top panel) and hospital admissions (bottom) are shown to compare the targeted calibration with the corresponding trajectory.

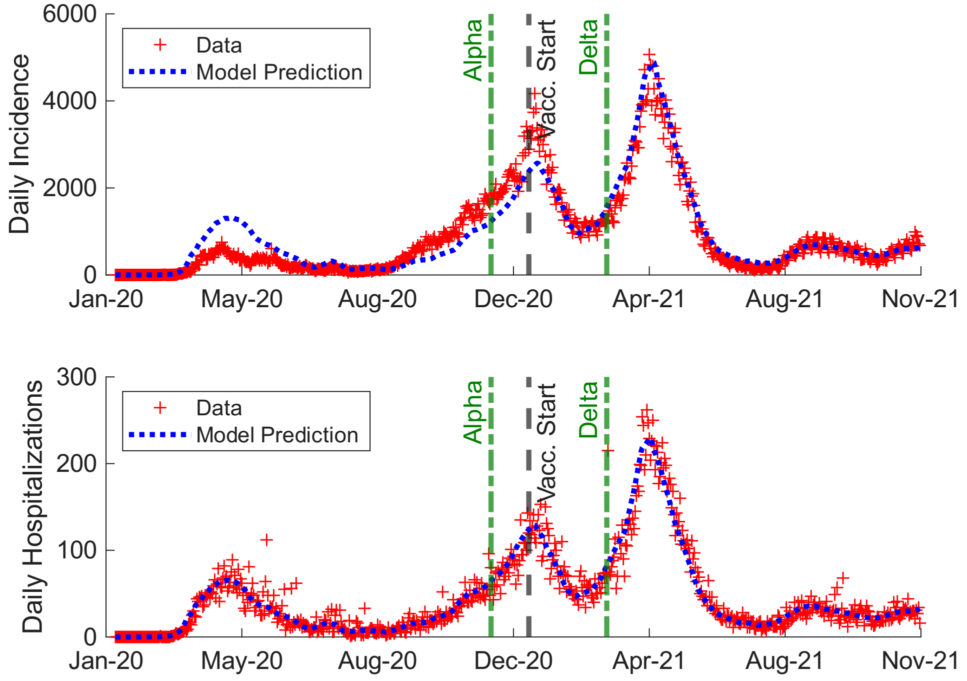


Figure S5: Model calibrated to hospital admissions only. Both incidence (top panel) and hospital admissions (bottom) are shown to compare the targeted calibration with the corresponding trajectory.

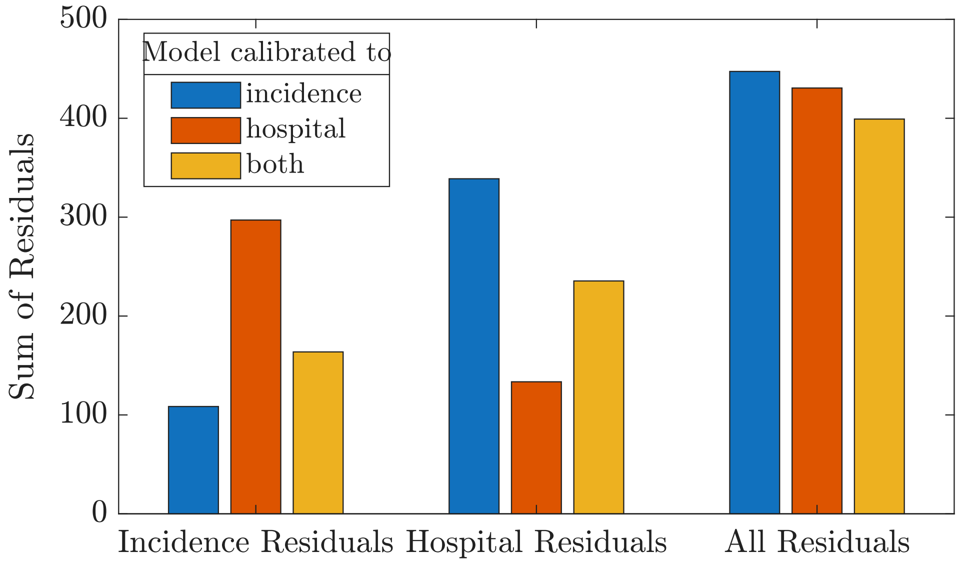


Figure S6: Comparison of residuals from models calibrated to incidence only, hospital admissions only, or both data streams. Bars show incidence, hospital, and total residuals.

#### S2.2 Counterfactual infections by severity

Figures S7–S9 show the counterfactual trajectories separately for mild, moderate, and severe infections. Although the absolute magnitudes differ across severity classes, the broad effects of the counterfactual interventions are consistent. In the vaccine–NPI crossover scenarios, removal of NPIs produces a large early epidemic wave, whereas scenarios without vaccination generate substantial later burden after the observed vaccine-rollout period. Removing both vaccination and NPIs produces particularly large infection burdens across all three severity classes.

The targeted NPI counterfactuals show a consistent setting-specific pattern across mild (Figure S7), moderate (Figure S8), and severe infections (Figure S9). Altering school restrictions produces the smallest changes relative to the observed scenario, workplace changes produce larger increases, and relaxing restrictions in other community settings produces the largest increases. Partial restrictions reduce the burden relative to complete opening, while changing the timing of school reopening shifts the corresponding infection peaks.

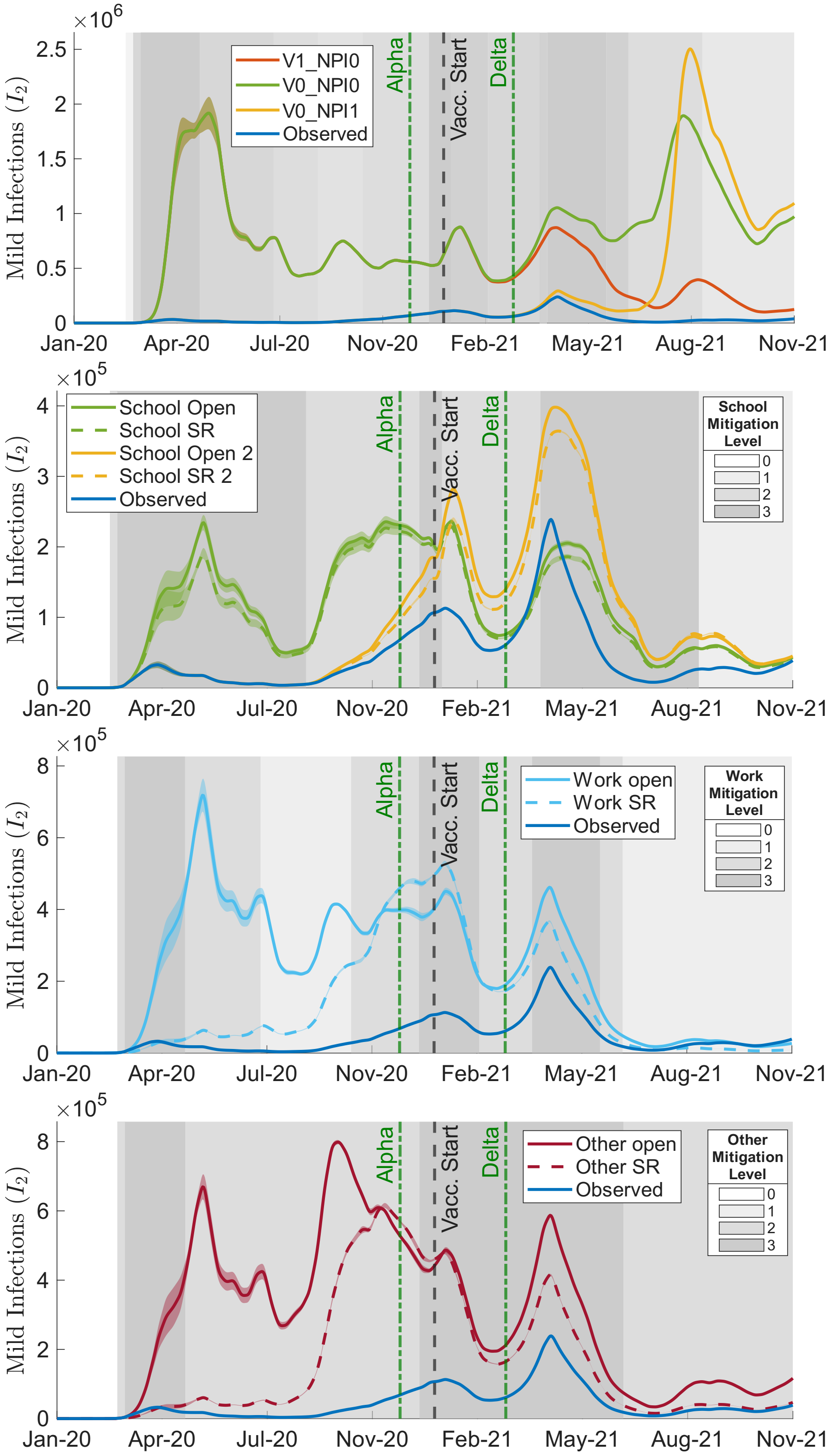


Figure S7: Counterfactual time series of mild infections $(I_2 + I_2^V)$. From top to bottom, panels show vaccine–NPI crossover scenarios, school-specific, workplace-specific, and other-setting NPI counterfactuals. Grey shading is based on NPI levels in the observed scenario and is included for timing context: the first panel shows the overall aggregate NPI level, while the remaining panels show the observed setting-specific NPI level for the corresponding school, workplace, or other setting. Darker shading indicates stricter measures.

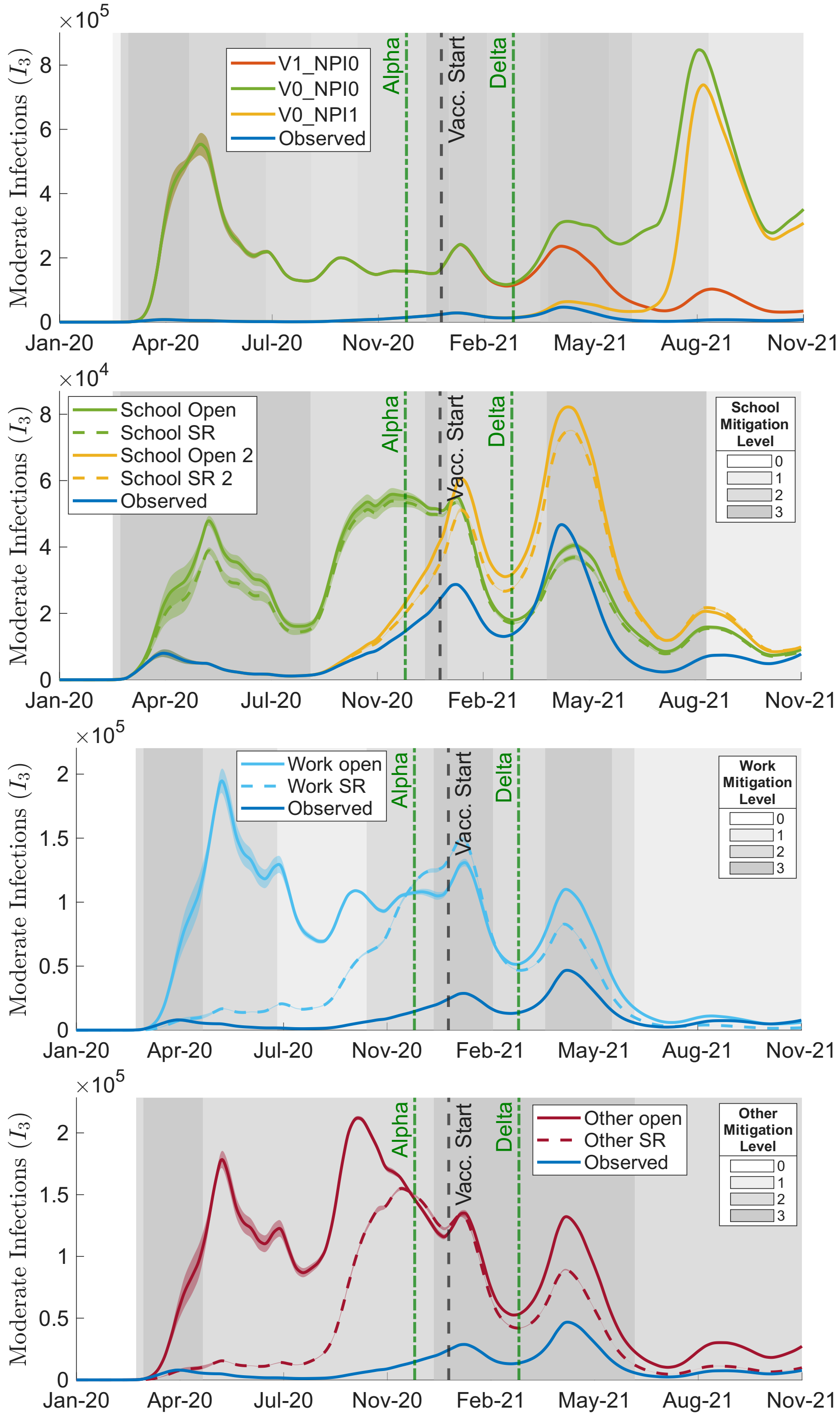


Figure S8: Counterfactual time series of moderate infections $(I_3 + I_3^V)$. From top to bottom, panels show vaccine–NPI crossover scenarios, school-specific, workplace-specific, and other-setting NPI counterfactuals. Grey shading is based on NPI levels in the observed scenario and is included for timing context: the first panel shows the overall aggregate NPI level, while the remaining panels show the observed setting-specific NPI level for the corresponding school, workplace, or other setting. Darker shading indicates stricter measures.

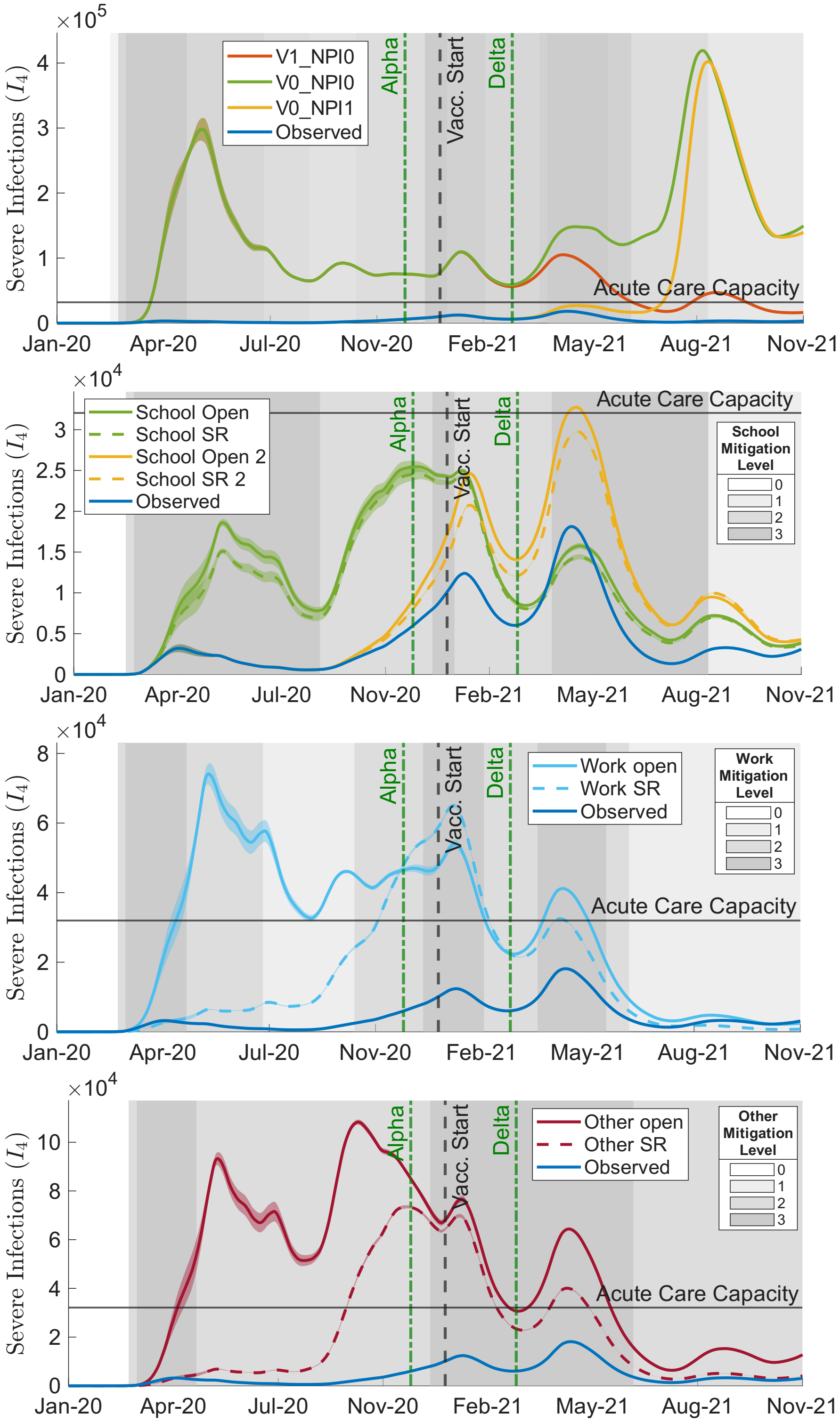


Figure S9: Counterfactual time series of severe infections $(I_4 + I_4^V)$. From top to bottom, panels show vaccine–NPI crossover scenarios, school-specific, workplace-specific, and other-setting NPI counterfactuals. Grey shading is based on NPI levels in the observed scenario and is included for timing context: the first panel shows the overall aggregate NPI level, while the remaining panels show the observed setting-specific NPI level for the corresponding school, workplace, or other setting. Darker shading indicates stricter measures.

#### *S2.3 Sensitivity analyses*

We conduct sensitivity analyses to assess the robustness of the model calibration and counterfactual conclusions to uncertainty in four key assumptions: the magnitude of school contact reductions under mitigation measures, the initial infected population, the rate of immunity waning, and the proportions used to map model-predicted daily incidence to reported case counts and hospital admissions.

##### *S2.3.1 Sensitivity to Calibration Parameters*

To compare model outputs with observed incidence and hospital-admission data, the baseline calibration assumes that 100% of incident severe infections, 60% of incident moderate infections, and 0% of incident mild infections are detected. Daily hospital admissions are estimated as 15% of daily severe incidence.

Because these quantities are uncertain, we examine alternative mappings from model-predicted daily incidence to surveillance data. The tested parameter sets independently vary severe-case detection, moderate-case detection, mild-case detection, and the fraction of daily severe incidence resulting in hospital admission (Table S5).

| Calibration Parameter Set | Severe detect | Moderate detect | Mild detect | Hosp / severe |
|---|---|---|---|---|
| B (Baseline) | 100% | 60% | 0% | 15% |
| S↓ (low severe detect) | **80%** | 60% | 0% | 15% |
| D↓ (low moderate detect) | 100% | **40%** | 0% | 15% |
| D↑ (high moderate detect) | 100% | **80%** | 0% | 15% |
| M5 (5% mild detect) | 100% | 60% | **5%** | 15% |
| M10 (10% mild detect) | 100% | 60% | **10%** | 15% |
| H↓ (low hosp) | 100% | 60% | 0% | **10%** |
| H↑ (high hosp) | 100% | 60% | 0% | **20%** |
| H↑↑ (very high hosp) | 100% | 60% | 0% | **25%** |

Table S5: Calibration parameter sets used in the sensitivity analysis. Columns specify the assumed detection proportions for daily severe, moderate, and mild incidence and the assumed fraction of daily severe incidence resulting in hospital admission.

For each calibration parameter set, we recalibrate the model and compare residuals for incidence and hospital admissions. The baseline parameter set provides the best overall balance between the two calibration targets (Figure S10). Alternative assumptions generally improve the calibration to one data stream at the expense of the other or increase the combined residual error, supporting the use of the baseline proportions in the main analysis.

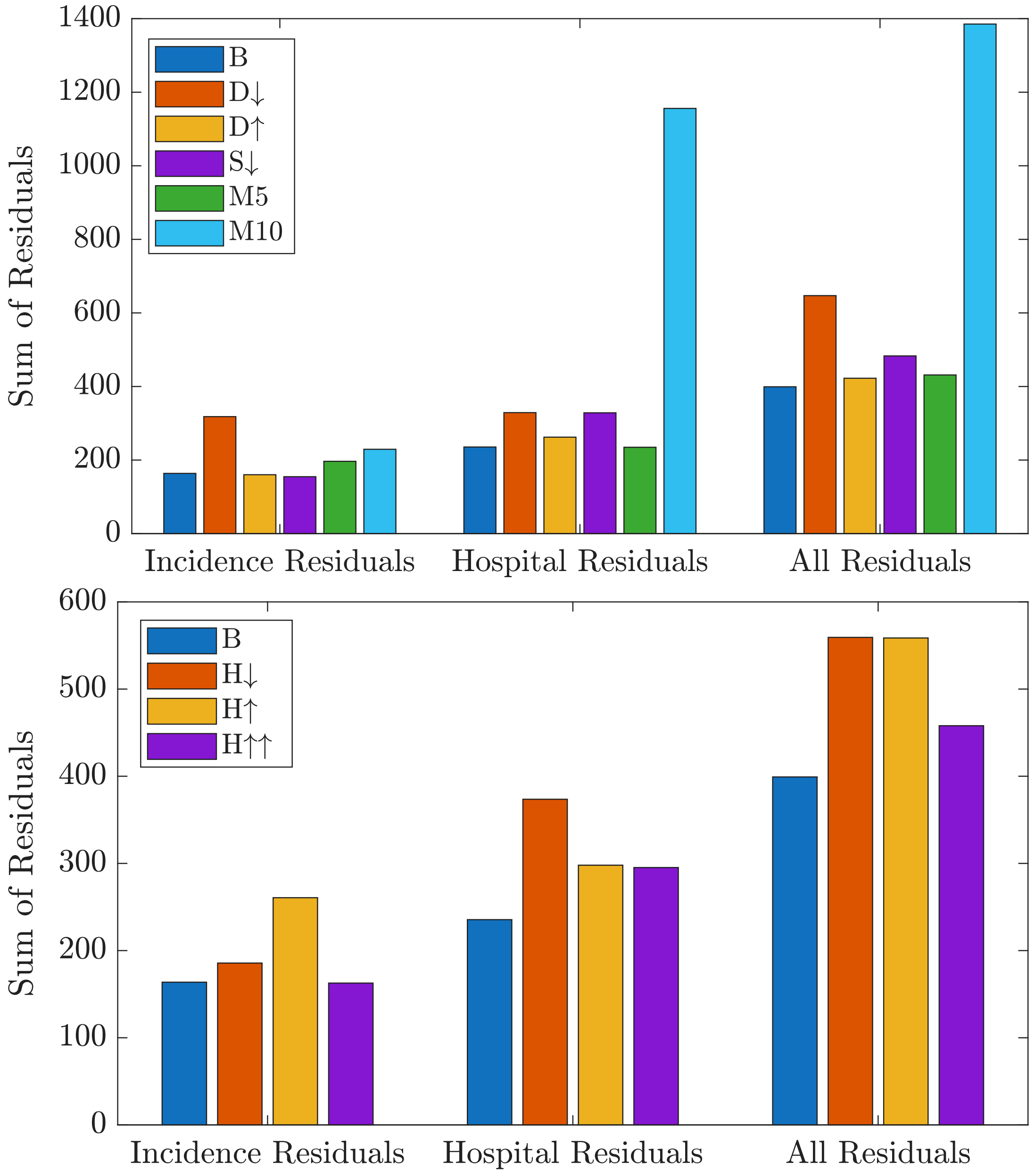


Figure S10: Sensitivity of calibration residuals to assumed detection and hospitalization proportions. Calibration parameter sets are defined in Table S5, and bars show incidence, hospital, and total residuals.

##### *S2.3.2 Sensitivity to School Contact Perturbation Assumptions*

The school perturbation matrix used in the main analysis assumes that on-site mitigation measures produce relatively modest reductions in school contacts. In particular, under mitigation level 1, child–child contacts are reduced by 5% relative to the baseline school contact matrix. Because the effectiveness of school-based measures such as masking, distancing, and enhanced hygiene is uncertain, we test alternative perturbation matrices representing progressively larger reductions in school contacts.

The alternative values are shown in Table S6. Values indicate the percentage of baseline school contacts remaining under each mitigation level. Contact rates corresponding to no restrictions and maximum closure are unchanged; the sensitivity analysis varies the contact multipliers associated with mitigation levels 1 and 2.

| Age Range | Mitigation Level | Baseline Values | Alternative 90-75-75-60 | Alternative 85-70-70-55 | Alternative 80-65-65-50 | Alternative 75-60-60-45 |
|---|---|---|---|---|---|---|
| 0–20 | 1 | **95%** | **90%** | **85%** | **80%** | **75%** |
| 20+ | 1 | **75%** | **75%** | **70%** | **65%** | **60%** |
| 0–20 | 2 | **85%** | **75%** | **70%** | **65%** | **60%** |
| 20+ | 2 | **75%** | **60%** | **55%** | **50%** | **45%** |

Table S6: School contact-rate multipliers used in the sensitivity analysis of school perturbation assumptions. Percentages indicate the remaining contact rate relative to the baseline school contact matrix for each age group and mitigation level.

For each alternative perturbation matrix, we repeat the school counterfactual simulations and compare the resulting hospital prevalence trajectories (Figure S11). Increasing the assumed effectiveness of school mitigation increases the separation between the School Open and School SR scenarios. However, the qualitative conclusion remains unchanged across the tested assumptions: the hospital prevalence generated by the school counterfactuals remains below the 25% acute-care-capacity reference level.

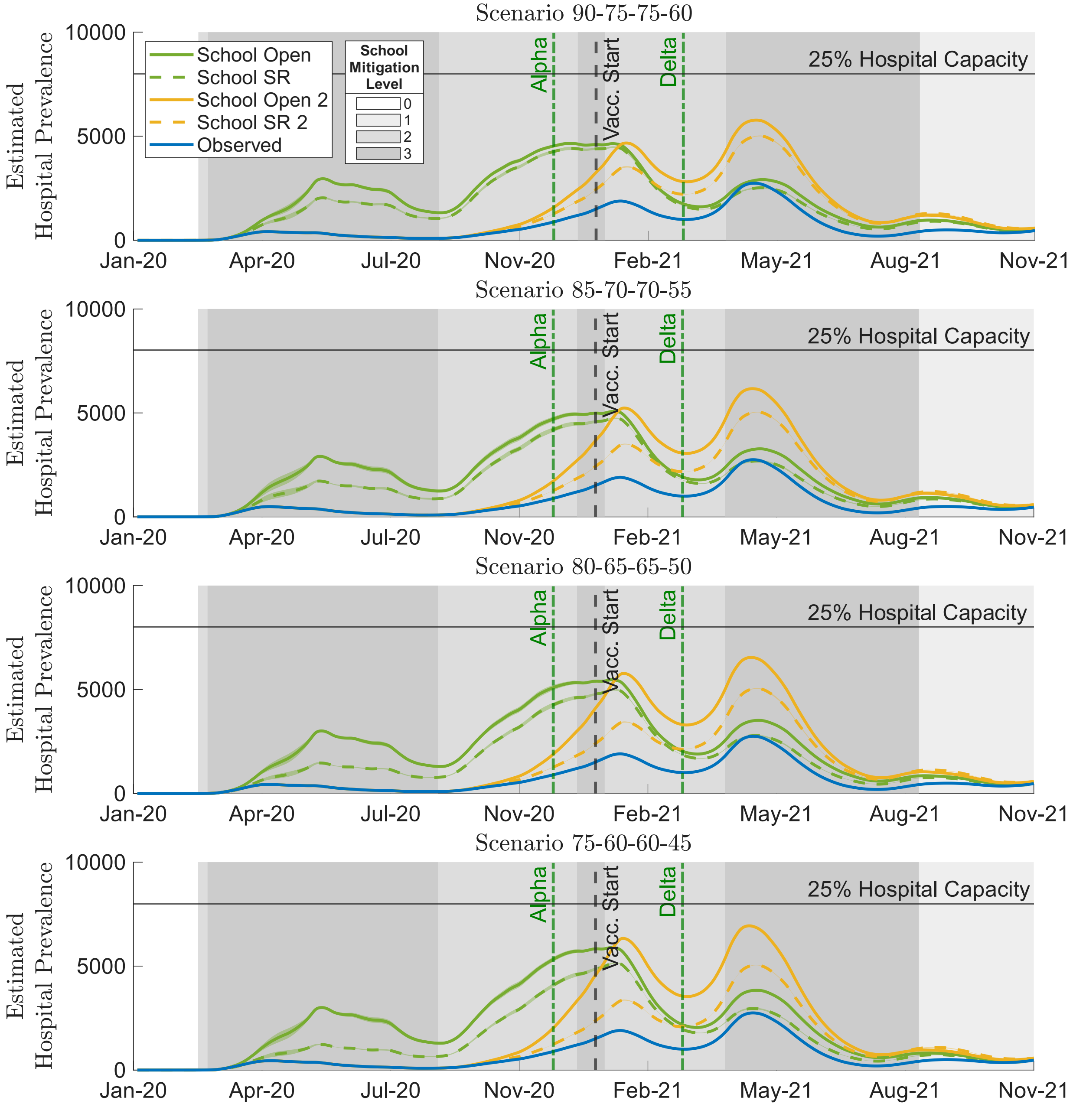


Figure S11: Sensitivity of school counterfactual hospital prevalence to alternative school contact-reduction assumptions. Panels correspond to the school perturbation sets listed in Table S6.

##### S2.3.3 Sensitivity to Initial Conditions

The baseline model is initialized by distributing a small infection seed across six age groups spanning ages 20–50, with $I_0 = 1/6$ assigned to each selected age group. To assess sensitivity to this assumption, we repeat the calibration using $I_0 = 0.1, 1/6, 0.5, 1, 1.5, 2,$ and 10.

The assumed initial infected population affects the calibrated transmission multiplier $\kappa(t)$ primarily during the first few weeks of the epidemic (Figure S12). Larger initial infection seeds require lower initial values of $\kappa(t)$, whereas smaller seeds require higher values. These differences diminish rapidly as calibration progresses, and the resulting counterfactual trajectories are nearly identical across the tested initial conditions.

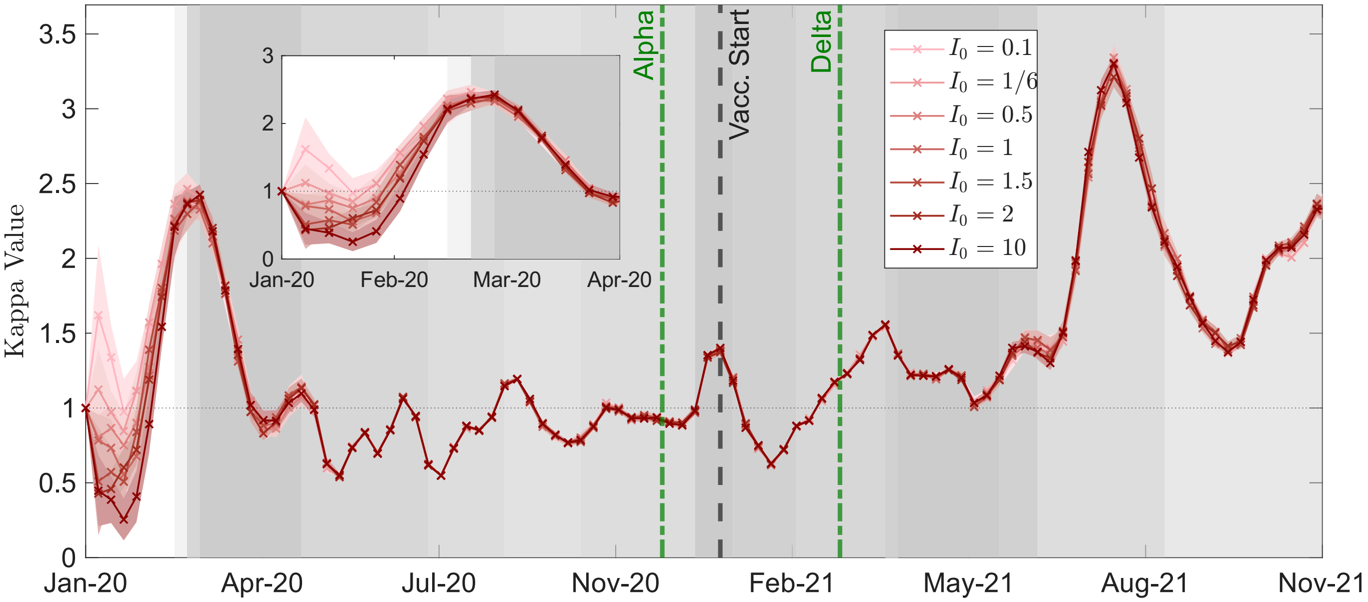


Figure S12: Sensitivity of calibrated $\kappa(t)$ to the assumed initial infected population $I_0$. The inset shows the first three months, where differences are largest.

##### S2.3.4 Sensitivity to Immunity Waning Rate

The baseline model assumes a one-year average duration before waning from each higher immunity class to the next lower class, corresponding to $\omega = 1$ year$^{-1}$ (equivalently, 1/365 day$^{-1}$). The same waning rate is applied across age groups and to unvaccinated susceptible, vaccinated susceptible, and vaccinated compartments.

To evaluate uncertainty in this assumption, we consider

$$\omega = \frac{1}{3}, \frac{2}{5}, \frac{1}{2}, \frac{2}{3}, 1, \frac{4}{3}, 2, \text{ and } 4 \quad \text{year}^{-1},$$

corresponding to average waning times of 3 years, 2.5 years, 2 years, 1.5 years, 1 year, 9 months, 6 months, and 3 months, respectively.

The calibrated $\kappa(t)$ trajectories remain similar across waning assumptions for most of the study period, although faster waning generally requires somewhat lower calibrated values of $\kappa(t)$, particularly later in the epidemic (Figure S13, top panel). Faster waning also increases hospital prevalence in the counterfactual simulations because individuals return to lower-immunity states more rapidly. Despite differences in magnitude, the relative behaviour of the observed and counterfactual scenarios remains qualitatively consistent across the tested waning rates (Figure S13).

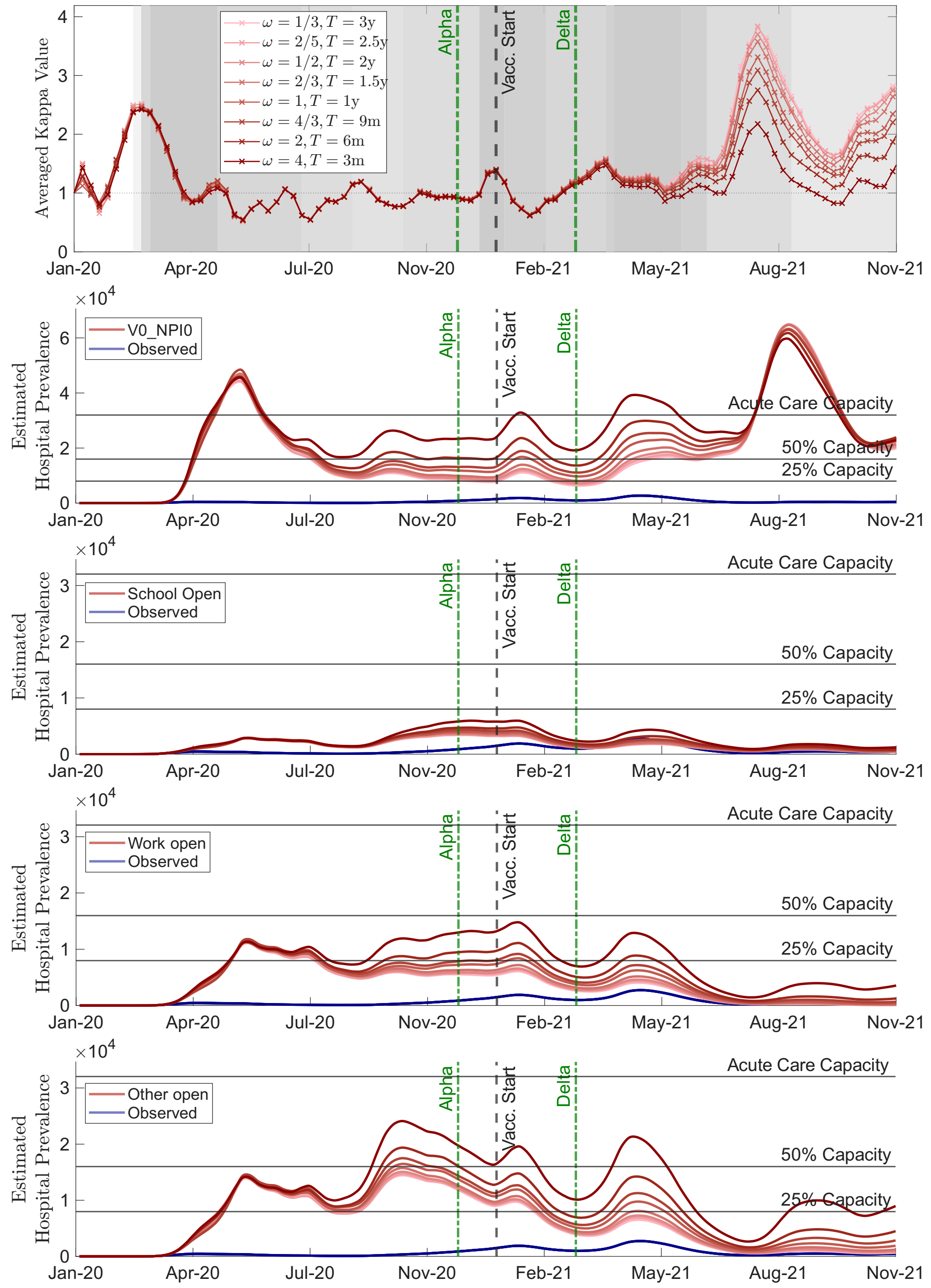


Figure S13: Sensitivity of model calibration and counterfactual hospital prevalence to the assumed immunity waning rate. From top to bottom: calibrated transmission multiplier $\kappa(t)$; no vaccination or NPIs; School Open; Work Open; and Other Open scenarios. The dark blue curve shows the baseline observed scenario, and red curves of different shades correspond to alternative waning rates. Here, $y$ and $m$ denote years and months, respectively. The baseline value $\omega = 1$ year$^{-1}$ ($T = 1$ year) corresponds to the value reported in Table S2.

#### S2.4 *Observed-scenario compartment dynamics*

The internal model trajectories under the observed scenario are shown in Figure S14. Before vaccine rollout, the population remains predominantly in the unvaccinated susceptible compartments, with temporary increases in infected compartments corresponding to the epidemic waves. Following vaccine introduction, the first-dose compartments increase rapidly and subsequently decline as individuals progress to second-dose compartments. Vaccinated-susceptible compartments become increasingly populated later in the study period as vaccine-derived immunity wanes, while vaccinated infections emerge after rollout but remain small relative to the susceptible and vaccinated population compartments. These trajectories illustrate how infection, vaccination, and waning jointly redistribute the population among the model's immunity states over the course of the observed epidemic.

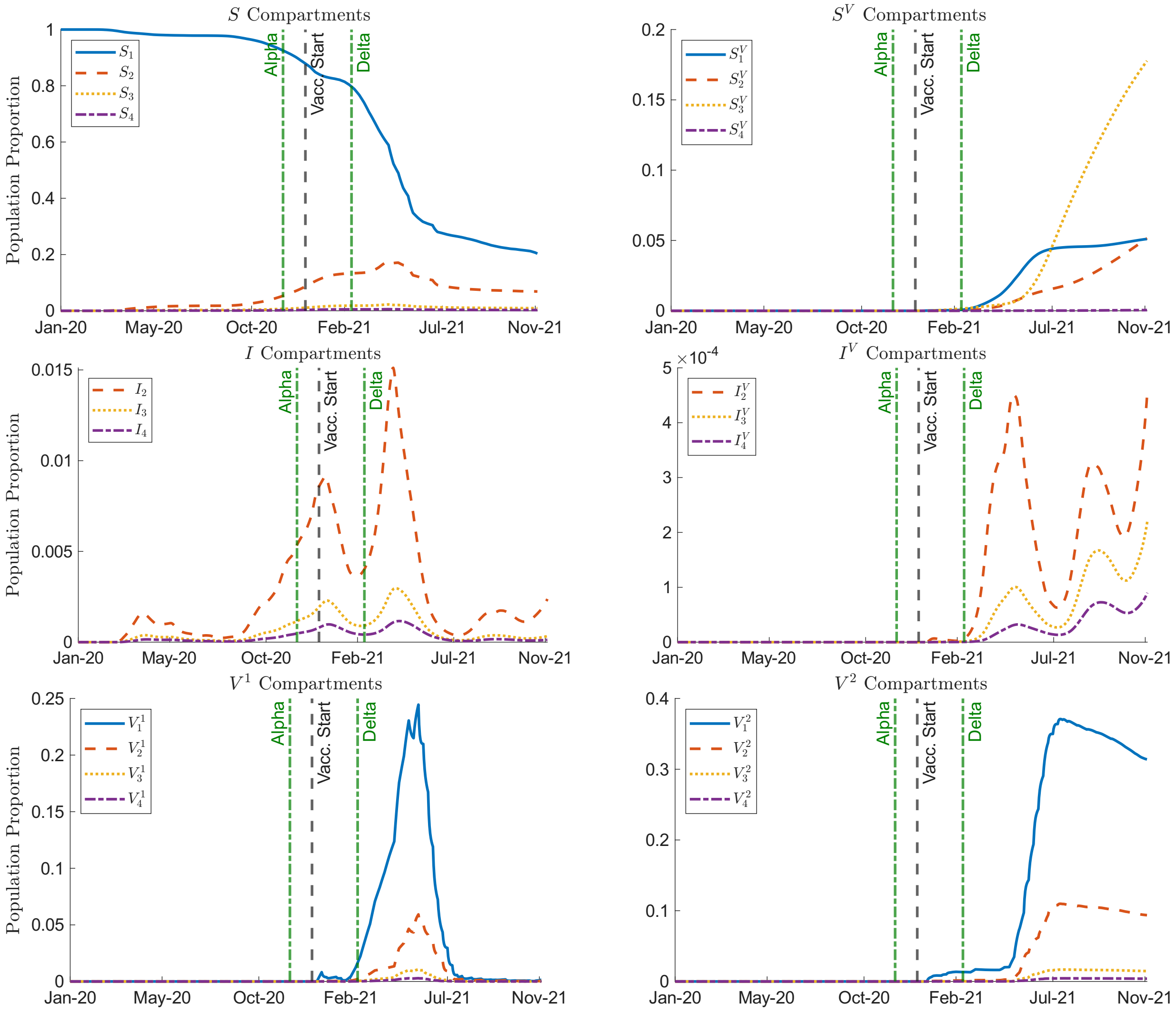


Figure S14: Time series of model compartment proportions under the Observed scenario. Panels show unvaccinated susceptible compartments $S_i$, vaccinated-susceptible compartments $S_i^V$, infected compartments $I_j$, vaccinated infected compartments $I_j^V$, first-dose vaccinated compartments $V_i^1$, and second-dose vaccinated compartments $V_i^2$. Within each susceptible, vaccinated-susceptible, and vaccinated panel, curves show the four immunity levels $i = 1, \ldots, 4$ separately; within infected panels, curves show symptom severity classes $j = 2, 3, 4$.

#### *S2.5 Wave-specific hospital burden*

The main counterfactual time series show that hospital-prevalence peaks do not occur at the same times across scenarios. Some counterfactuals generate large early waves, whereas others shift the largest burden to the Alpha, Delta, or post-Delta periods. To compare scenarios more consistently, we divide the pre-Omicron period into fixed wave windows: Wave 1, Pre-Alpha, Alpha, Delta, and Post-Delta. Within each window, we compute the maximum hospital prevalence and the peak 6-week mean, both expressed as percentages of Ontario acute-care capacity (Supplementary Figure S15). The 6-week mean summarizes sustained hospital pressure around the peak, whereas the maximum captures the largest instantaneous burden.

For the vaccine–NPI crossover scenarios (left panels), removal of NPIs dominates the first wave. The `V1_NPI0` scenario produces a large first-wave hospital-prevalence peak, while the observed trajectory remains near zero relative to acute-care capacity. Because vaccination has not yet begun, the `V0` scenarios are not shown separately in the first two wave windows. In the Pre-Alpha period, `V1_NPI0` again generates a substantially higher peak than the observed scenario, indicating that NPIs are clearly the dominant control mechanism before vaccination can affect population-level hospital burden.

During the Alpha window, the `V1_NPI0` and `V0_NPI0` scenarios produce the largest crossover burdens, both reaching approximately the 50% acute-care-capacity reference level. The `V0_NPI1` scenario is much closer to the observed trajectory during this period, suggesting that NPIs still provided substantial protection before vaccine-derived immunity was widespread. In the Delta window, the absence of both vaccination and NPIs produces the largest peak, while `V1_NPI0` and `V0_NPI1` are lower but still elevated relative to the observed scenario. By the Post-Delta period, the effect of vaccination becomes especially clear: scenarios without vaccination, particularly `V0_NPI0` and `V0_NPI1`, produce the largest sustained hospital burden, approaching or exceeding very high fractions of acute-care capacity.

The targeted NPI adjustments (right panels) show a consistent hierarchy across wave windows. School-only changes produce the smallest hospital-prevalence increases. Even when schools are fully open, the wave-specific peaks remain below the 25% acute-care-capacity reference line. Opening schools only in the second academic year shifts more burden into the Alpha and Delta windows, but the resulting peaks remain much smaller than those generated by workplace or other-setting relaxation.

Workplace reopening produces larger and more sustained increases than school reopening. Work Open generates a pronounced first-wave burden and remains elevated through the Pre-Alpha, Alpha, and Delta periods. Work SR reduces this burden relative to full reopening, but still produces peaks above the observed trajectory, especially around the Alpha period. These results indicate that even partial workplace restrictions substantially reduce hospital pressure, but do not fully reproduce the protection achieved by the observed layered intervention strategy.

Relaxing restrictions in other community settings produces the largest targeted-NPI effects. Other Open generates high hospital-prevalence peaks in the first wave, Pre-Alpha, Alpha, and Delta windows, with the Pre-Alpha period showing the largest sustained burden among the targeted scenarios. Other SR reduces hospital prevalence relative to Other Open, but still produces substantial peaks, particularly in the Pre-Alpha and Alpha periods.

Overall, the wave-specific summaries confirm the conclusions from the full time-series plots while clarifying when each intervention matters most. NPIs are most important before vaccination and during the early variant periods, whereas vaccination becomes most important in preventing the large post-Delta burden. Among targeted NPIs, other-setting restrictions contribute the most to controlling hospital prevalence, workplace restrictions have an intermediate effect, and school restrictions have comparatively modest effects on severe healthcare demand.

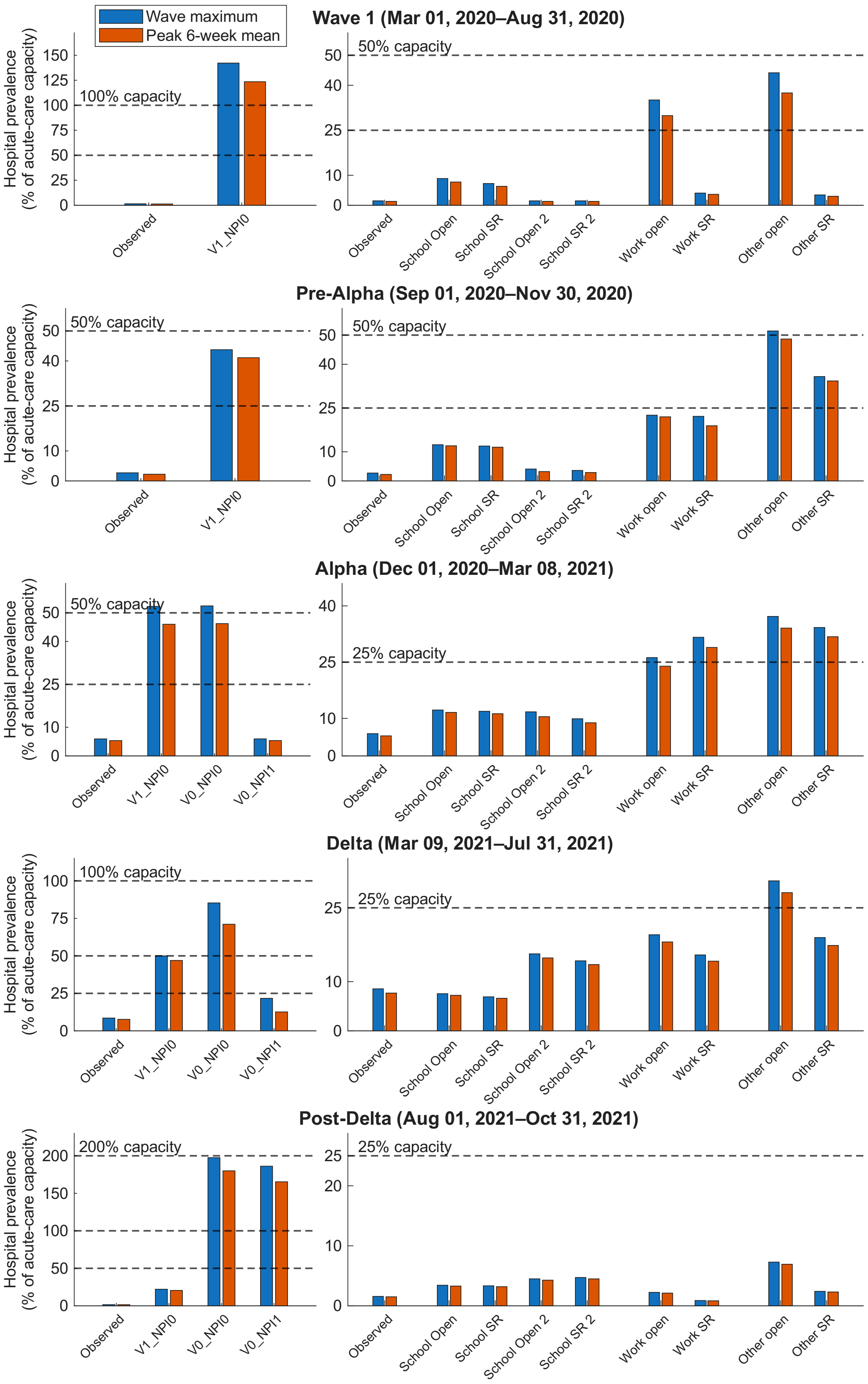


Figure S15: Ontario wave-specific hospital-prevalence summaries. Rows show Wave 1, Pre-Alpha, Alpha, Delta, and Post-Delta; left and right panels compare vaccine–NPI crossovers and targeted NPI adjustments, respectively. Bars show wave maxima and peak 6-week means as percentages of acute-care capacity.

#### *S2.6 Cumulative counterfactual outcomes by severity*

The cumulative results reinforce the patterns observed in the time-series analyses. Across mild, moderate, and severe infections, removing both vaccination and NPIs produces the largest absolute and relative increases compared with the observed scenario (Figure S16). Among the setting-specific counterfactuals, relaxing restrictions in other community settings produces the largest cumulative increase, followed by workplace relaxation, whereas school-specific changes produce substantially smaller increases. For each setting, retaining some restrictions reduces the cumulative burden relative to complete opening.

These severity-specific results are consistent with the combined total-infection and hospital-admission outcomes presented in Figure 6 of the main manuscript.

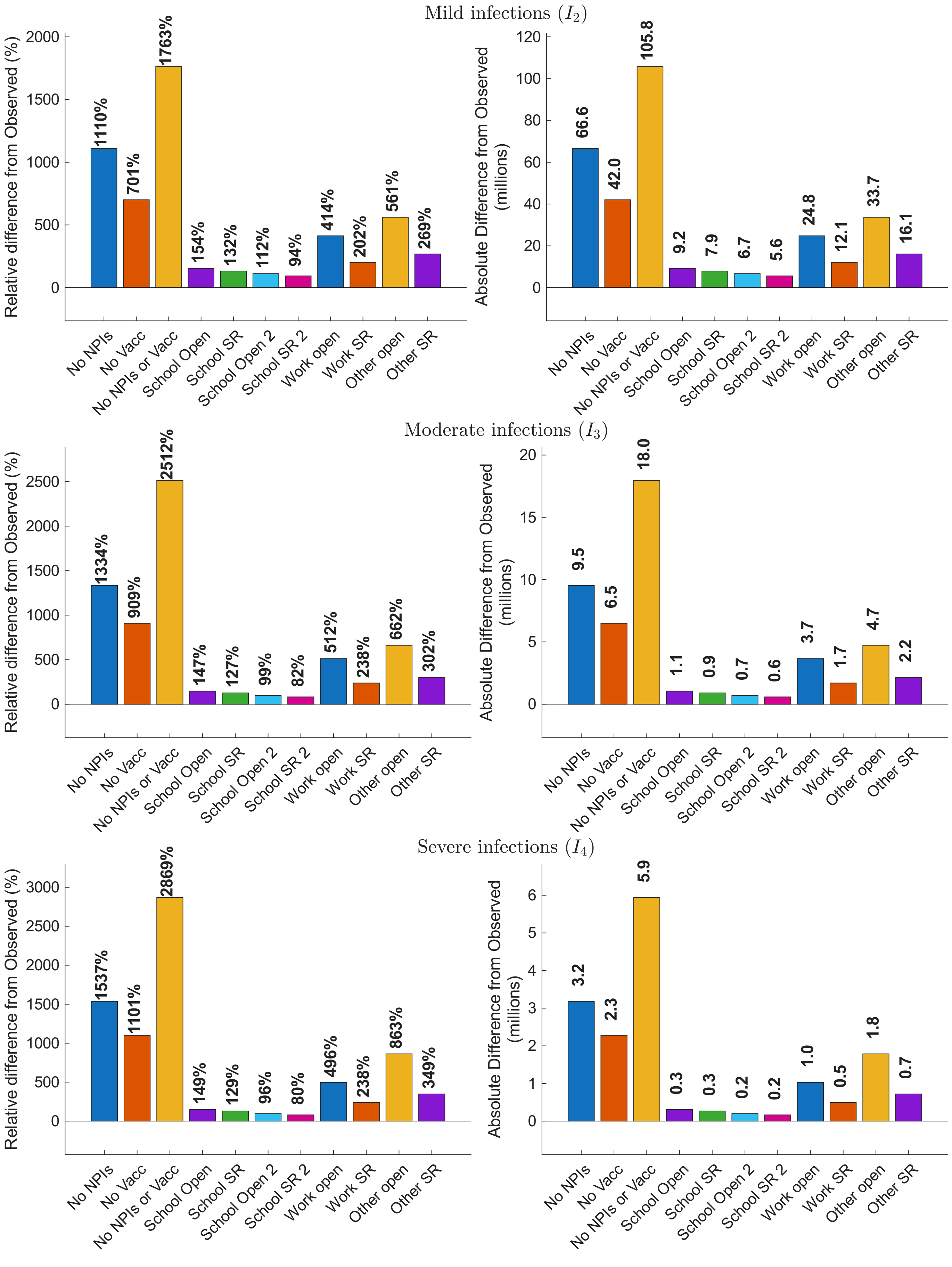


Figure S16: Differences in cumulative infections by severity class compared with the Observed scenario. Rows show cumulative mild, moderate, and severe infections, from top to bottom. In each row, the left panel shows the relative difference from the Observed scenario, and the right panel shows the absolute difference.

#### *S2.7 Additional immunity distributions*

Figure S17 extends the immunity analysis in the main manuscript to the remaining vaccine–NPI crossover scenarios. In the `V1_NPI0` scenario, the absence of NPIs allows substantial infection-derived immunity to accumulate before vaccine rollout; vaccination subsequently shifts an increasing fraction of the population toward higher immunity classes. In contrast, under `V0_NPI1`, NPIs preserve a larger low-immunity population for longer, but in the absence of vaccination, later immunity gains arise primarily through infection.

The targeted Work Open and Other Open scenarios are shown in Figure S18. Both produce greater pre-vaccine movement away from the lowest immunity class than the observed scenario, with the shift occurring more strongly under Other Open, consistent with its larger infection burden. Following vaccine rollout, both scenarios show substantial movement into higher immunity classes. Together with the scenarios shown in the main manuscript, these results illustrate that similar levels of population immunity can arise through very different pathways: vaccination can increase immunity while limiting infection burden, whereas relaxed NPIs generate additional immunity largely through infection.

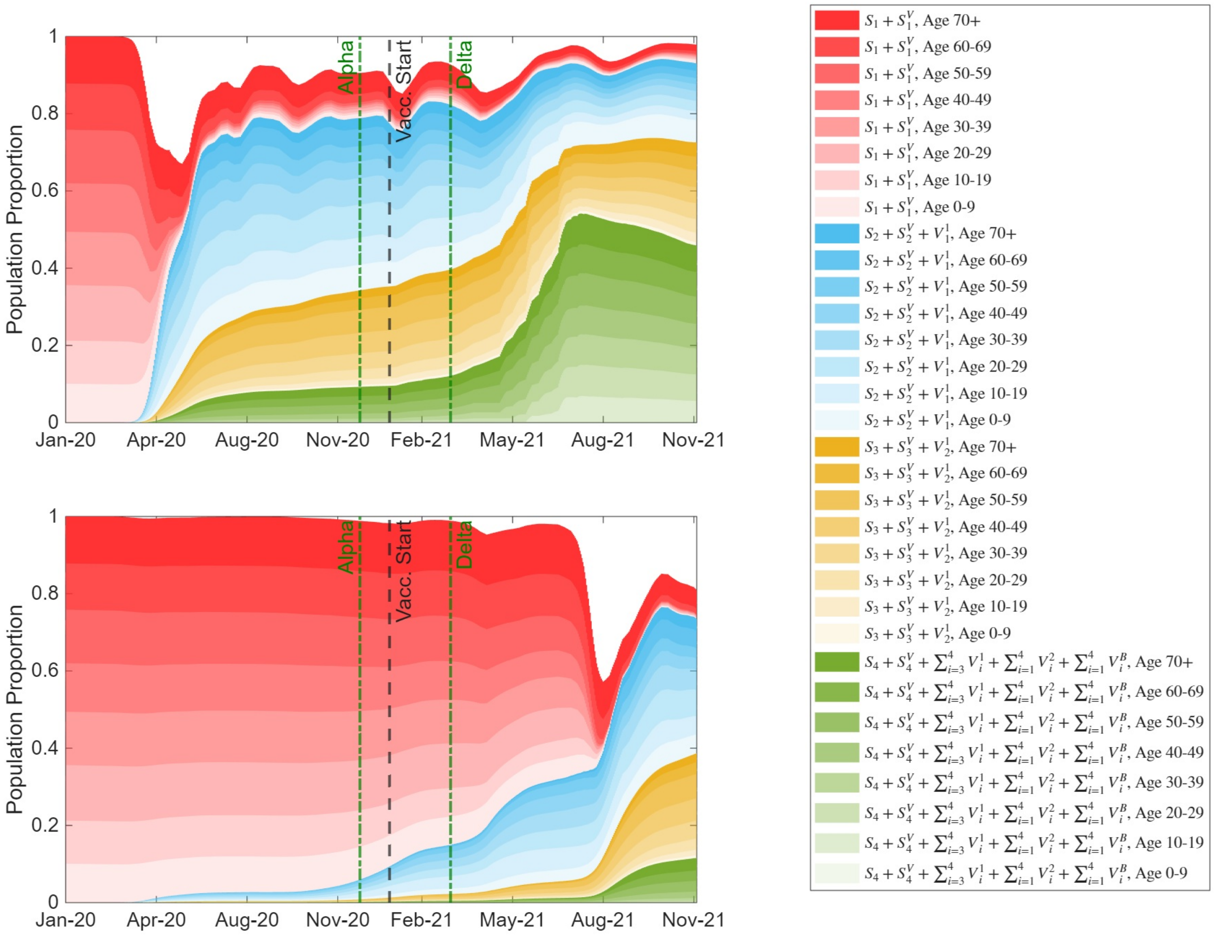


Figure S17: Immunity distribution by age group and immunity class in the `V1_NPI0` and `V0_NPI1` scenarios. Colours and white space are defined as in Figure 7 in the main manuscript.

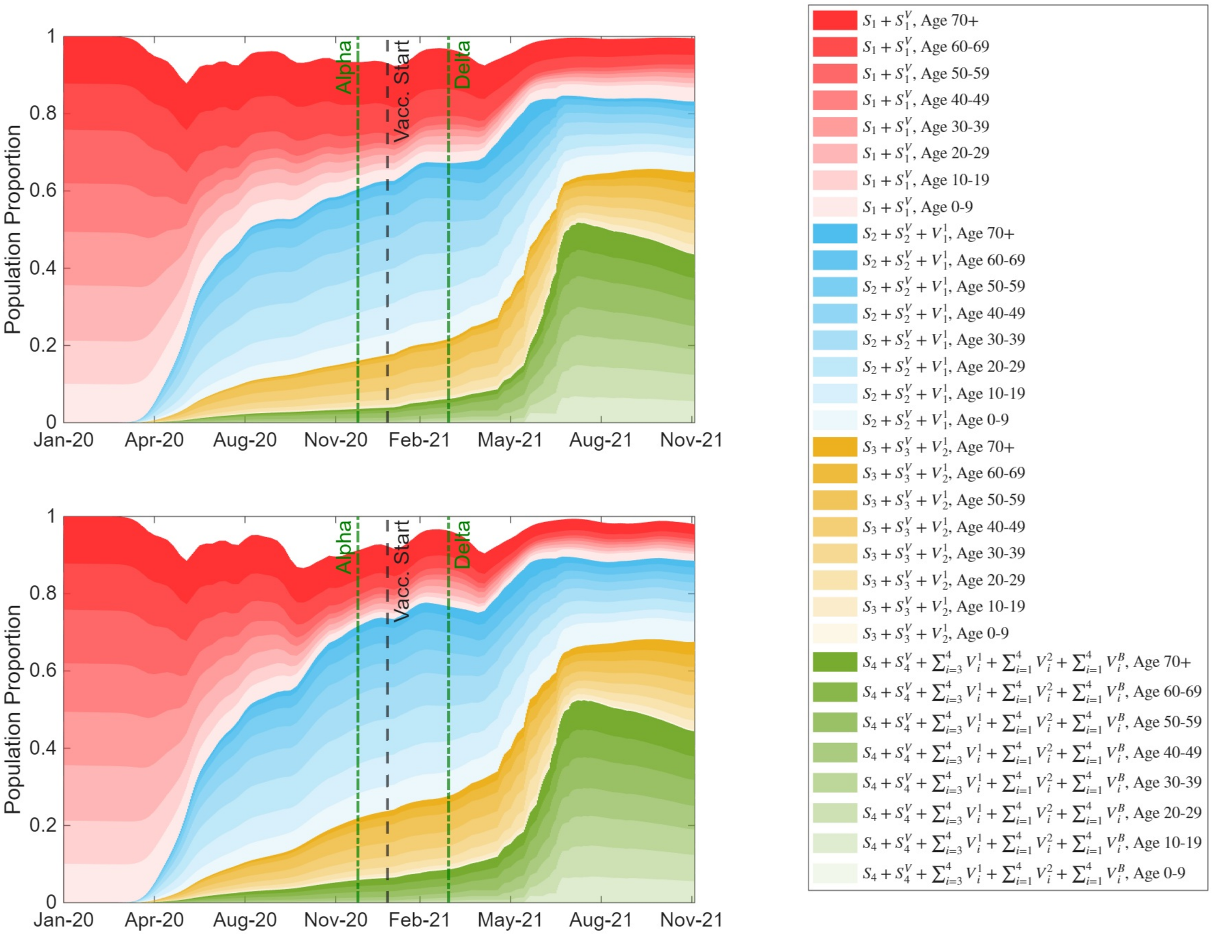


Figure S18: Immunity distribution by age group and immunity class in the Work Open and Other Open scenarios. Colours and white space are defined as in Figure 7 in the main manuscript.

#### S2.8 Comparison with Canada-wide counterfactuals

We compare our Ontario counterfactual projections with the Canada-wide agent-based modelling results reported by Ogden et al. [24]. The authors simulated a representative Canadian population of 100,000 agents and reported cumulative clinical cases, asymptomatic cases, hospitalizations, ICU admissions, and deaths per 100,000 population for several counterfactual scenarios from February 7, 2020 to March 31, 2022. In contrast, our deterministic model represents the full Ontario population used in our calibration. Although the per-capita outputs from Ogden et al. can be scaled to Ontario for contextual comparison, their 100,000-agent simulation remains a representative national population rather than an Ontario-specific full-population projection. This difference in population representation, together with stochasticity and household/contact clustering in the agent-based model, may contribute to differences in cumulative infections after scaling, particularly compared with a deterministic population-level model in which transmission is averaged over the full Ontario population.

To make the results comparable in scale, we multiply the per-capita estimates from Ogden et al. by the Ontario population used in our model. We then match their clinical cases to our cumulative moderate and severe infections, asymptomatic cases to our cumulative mild infections, total infections to the cumulative sum across all severity classes, and hospitalizations to our cumulative hospital admissions (Figure S19). Our model generally predicts larger cumulative infection totals than the scaled estimates from Ogden et al., particularly for asymptomatic and total infections (Figure S19). This occurs even though Ogden et al. include the Omicron wave, whereas our comparison excludes it. The discrepancy should therefore not be interpreted as a simple consequence of variant inclusion. Instead, it likely reflects differences in geographic scope, population representation, contact structure, public health feedback, symptom assumptions, and immunity representation.

First, Ogden et al. simulate a Canada-wide epidemic in a representative Canadian population, whereas our model is calibrated specifically to Ontario. National per-capita estimates average over substantial regional variation in demography, contact rates, epidemic intensity, and public health responses. This is important because the epidemic experience differed markedly across Canada, with some jurisdictions maintaining very low transmission for extended periods while larger and more urbanized provinces experienced repeated waves. Scaling Canada-wide estimates to the Ontario population therefore does not necessarily reproduce an Ontario-specific counterfactual.

Second, model structure may contribute to the difference. Our deterministic framework uses age-structured population-level mixing, whereas the agent-based model used by Ogden et al. includes household and venue structure. Household clustering can generate repeated contacts and local susceptible depletion, which may reduce transmission efficiency relative to a more population-level mixing approximation. In addition, the finite 100,000-agent structure of their model represents a stochastic sample of the Canadian population, while our model averages transmission over the full Ontario population. These structural differences can influence cumulative infections, especially under high-transmission counterfactuals.

Third, Ogden et al. explicitly represent several public health response mechanisms, including case detection, isolation, contact tracing, and quarantine. These mechanisms can remove infectious or exposed individuals from circulation, particularly as incidence increases. By contrast, our counterfactuals retain the calibrated transmission multiplier $\kappa(t)$ and do not fully represent incidence-triggered feedback through intensified testing, isolation, quarantine, or voluntary behavioural change. As a result, infections in high-transmission counterfactuals may grow more freely in our model than in an agent-based model where rising incidence can trigger additional individual- or system-level responses.

Fourth, differences in symptom classification may affect both transmission and the clinical/asymptomatic split. Ogden et al. assign a substantial fraction of infections in children and youth to symptomatic disease, whereas our clinical progression assumptions make symptomatic infection uncommon in younger age groups. If pediatric infections are more often asymptomatic or mildly symptomatic, symptom-based detection and isolation would miss a larger fraction of infections, allowing more silent onward transmission [75–77]. This difference may partly explain why our model predicts substantially more asymptomatic infections and total infections in several scenarios.

Finally, both models include waning immunity, but they implement it differently. In our model, waning begins immediately from each immune state at a specified transition rate, so individuals continuously move from higher to lower immunity levels over time. By contrast, Ogden et al. assume an initial period of preserved protection before waning begins: in their agent-based model, waning starts after a 90-day period following full recovery from infection or following receipt of a second or booster vaccine dose. This delay creates a period during which recently infected or vaccinated individuals remain more strongly protected before returning toward susceptibility. In contrast, our continuous waning structure allows immunity to decline from the time individuals enter each immune class. Differences in the timing of waning onset, waning rates, immune-state structure, and the way partially protected individuals return to susceptibility can therefore affect cumulative infections.

Overall, this comparison should be interpreted as qualitative rather than directly quantitative. Both models support the same broad conclusion that removing public health measures and/or vaccination would produce substantially larger infection and hospitalization burdens. Although our model generally predicts larger cumulative infection totals, especially for asymptomatic and total infections, the differences in hospital admissions are less pronounced, as shown in the fourth panel of Figure S19. This suggests that some of the discrepancy is driven by differences in infection classification, transmission structure, and asymptomatic spread rather than a uniformly larger predicted burden across all outcomes. The larger infection estimates in our model likely reflect Ontario-specific calibration, full-population deterministic transmission, more efficient population-level mixing, limited incidence-triggered behavioural feedback, differences in symptom classification, and differences in immunity structure, rather than disagreement about the protective effects of vaccination and public health measures.

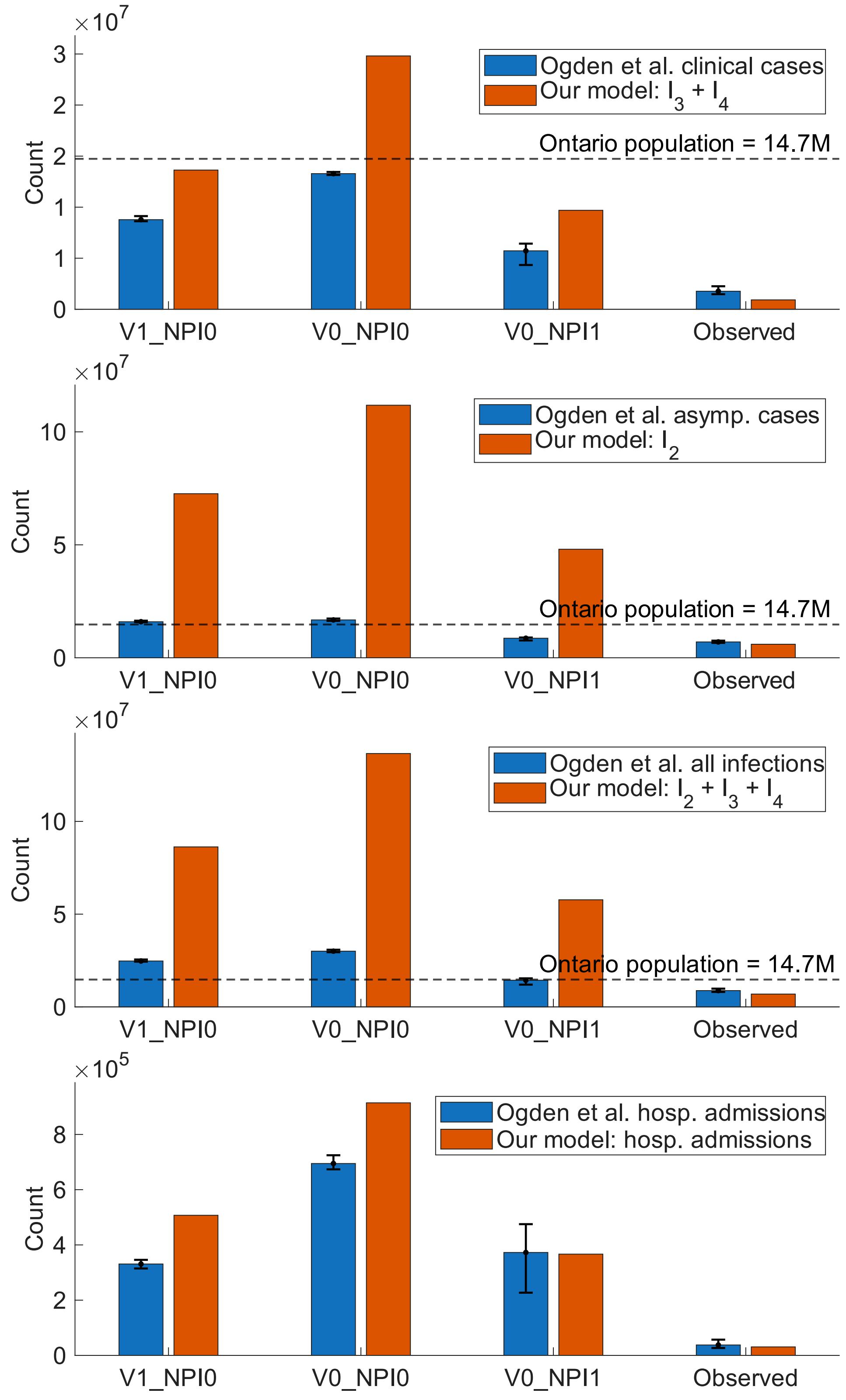


Figure S19: Comparison of our Ontario model outputs with the Canada-wide counterfactual estimates of [24] scaled to the Ontario population. Panels compare clinical cases, asymptomatic cases, total infections, and hospital admissions across matched scenarios.
